# Measuring Social Determinants of Health in the *All of Us* Research Program: Technical Document

**DOI:** 10.1101/2023.06.01.23290404

**Authors:** Samantha Tesfaye, Robert Cronin, Maria Lopez-Class, Qingxia Chen, Christopher S. Foster, Callie Gu, Andrew Guide, Robert A. Hiatt, Angelica Johnson, Christine LM Joseph, Parinda Khatri, Sokny Lim, Tamara R. Litwin, Fatima A. Munoz, Andrea Ramirez, Heather Sansbury, David G. Schlundt, Emma N. Viera, Elif Dede-Yildirim, Cheryl R. Clark

## Abstract

**Background:** To advance precision medicine and improve population health, the *All of Us* Research Program aims to collect data, including a survey of social determinants of health (SDOH), from over 1 million participants. This report (1) outlines the process used to construct the *All of Us* SDOH survey and (2) presents the psychometric characteristics and scoring recommendations for SDOH survey measures.

**Methods:** A consensus process was used to select a definition of SDOH and conceptual frameworks to develop inclusion and exclusion criteria and priorities for construct consideration. Cognitive interviewing was used to provide an assessment of SDOH survey item performance in English and Spanish.

Scales and scored items were constructed in alignment with validated literature. Item non-response was calculated, and Cronbach ‘s alpha was used to analyze the psychometric properties of each scale, overall and by demographic characteristics. Multivariable logistic regression models were used to examine if demographic categories were associated with item non-response.

**Results:** Twenty-nine percent (N=117,783) of *All of Us* participants submitted SDOH survey data by June 30, 2022. Among those who provided any SDOH survey data, item non-response was infrequent, with most scales having less than 5% incalculable scores due to item non-response. Item non-response varied most along the lines of racial identity, educational attainment, and the language in which the survey was administered (Spanish or English). In our regression models, for most scales, patterns of missing data due to item non-response were seen by racial identity, educational attainment, income level, and age. Internal consistency reliability was greater than 0.80 for almost all scales, with variability by racial identity, educational attainment, and the language of survey administration.

**Conclusion:** The SDOH survey demonstrated good to excellent reliability across several measures of SDOHs and within multiple population groups that are underrepresented in biomedical research. Bias due to survey non-response and item non-response should be monitored and addressed as the survey is fielded more completely.

## Introduction

There is growing scientific consensus that social factors are significant contributors to health status, including the onset or progression of disease, recovery or response to treatment, and inequities in distributions of illnesses among populations.^1,2^ Investigating the roles of social factors in health is an essential part of precision medicine research.

The *All of Us* Research Program (*All of Us*) aims to collect data from over 1 million participants to advance precision medicine and improve population health.^3^ The program seeks to build a diverse cohort that recruits populations underrepresented in biomedical research (UBR) along the lines of race, ethnicity, age, sex, gender identity, sexual orientation, disability status, access to care, income, educational attainment, and geographic location.^3,4^ A central goal of *All of Us* is to ensure program participants represent the diversity of the US. *All of Us* supports research that integrates information on genomics and other biologic data, data on lifestyle or behaviors, and contextual factors including the social determinants of health (SDOH) to facilitate precision medicine research. Data from *All of Us* participants are captured in multiple ways, including health surveys to obtain participant provided information, electronic health record data, physical measurements, and biospecimens.

To collect data on social factors, *All of Us* launched a Task Force of subject matter experts within the *All of Us* Research Program to develop a survey on SDOH. SDOH concepts were selected to provide psychometrically rigorous data with strong use cases relevant to precision medicine. This technical report outlines the approach and process used to construct the SDOH survey. Additionally, this report presents psychometric characteristics of the measures included in the final SDOH survey, based on the most recently released data from 117,783 *All of Us* participants who completed any portion of the survey by June 30, 2022.

## Methods

### Survey development process

In 2017, the SDOH Task Force launched for the purpose of developing a survey to gather self-reported data from *All of Us* participants that captures information on dimensions of SDOH using scientifically valid and reliable scales, while minimizing burden on participants to complete surveys.

A six-phase process was developed by the Task Force to select concepts that could be measured with validity and reliability via self-report in a large, diverse, and multilingual cohort. This process is summarized in Appendix 1. In brief, the Task Force employed a consensus based process to define SDOH and conceptual frameworks to (1) establish the inclusion and exclusion criteria, and priorities to guide construct selection; (2) evaluate relevant scientific literature to identify measures with strong psychometric properties and use cases in precision medicine; and (3) examine SDOH measures used in other biobanks and large epidemiologic studies to find opportunities to align measures with other resources, chiefly the UK Biobank, the Million Veteran Program, and the NIH PhenX Toolkit surveys related to SDOH.^5-7^ Additionally, the Task Force coordinated internally to (4) avoid duplication of other *All of Us* surveys already deployed or in development (e.g., mental health, environmental/occupational exposures, and a survey offered to all participants as they join the program called “The Basics”) that assess concepts that overlap with or may be considered SDOH (e.g., income, education, employment, health literacy, home ownership, and risk of homelessness).^8^ The Task Force used conceptual frameworks to improve the user experience and eliminated lengthy scales that added to participant burden. Final measurement selections were completed with recommendations from scientific subject matter experts and *All of Us* participant partners. Importantly, inclusion and exclusion criteria for surveys prioritized measures of *perceptions* (including cognitions, beliefs, attitudes) that could only be obtained from the perspective of participants, and not otherwise collected through geocoding, electronic health records, or other means. Priority was given to selecting constructs and measures that are likely to represent core drivers and mechanisms connected to health inequities.^9,10^

The final survey included concepts and measures that had been previously validated and that had documentation on psychometric performance in large cohort studies. However, in rare instances, measures or items were included where extant literature is emerging when domain areas were considered essential for measurement, namely housing instability and housing quality.^11^ Priority was given to measures that were scientifically validated in multiple languages and cultural contexts. In general, measures were used in the manner in which they were validated with their original response sets. A notable exception was made for the questions on spirituality and religious service attendance, where a “non-religious” response option was added in keeping with strong recommendations from *All of Us* participants to acknowledge differing views on religion.

### Conceptual frameworks and definitions

The SDOH Task Force chose the World Health Organization Conceptual Framework for Action on the Social Determinants of Health as an organizing scientific framework to identify constructs that could be operationalized to establish connections among SDOHs, their origins in structural social, economic, political, historical and cultural factors, measures of social identity and social stratification (income, education, race, ethnicity, sexual orientation and gender identity, disability) and relation to psychological, material resources/health-related social needs, and social connections that influence health or health care utilization.^1^ Additionally, the Task Force sought to align with the Healthy People 2020 framework as described by the Centers for Disease Control and Prevention (CDC) Office of Disease Prevention and Health Promotion to identify five broad domains to refine the measurement approach: *social and community context, economic stability, education, neighborhood and built environment, health and health care*.^12^ Of these domains, the Task Force identified educational attainment and health care access as domains that were, in part, already covered in other *All of Us* surveys.

To improve the user experience and ability to communicate well with large, diverse audiences about the relation of social factors to health, the Task Force was guided by the work of the Robert Wood Johnson Foundation Commission to Build a Healthier America, which developed language and concepts to communicate about social factors across political groups.^13^

Finally, the Task Force solicited guidance and recommendations from *All of Us* Participant Ambassadors on concepts and specific measures resonant with, or that might be less relevant to, lived experience of participants.^14^ The Task Force also reviewed measures with scientific subject matter experts who were the developers of measurement tools under consideration, as well as subject matter experts from institutes and centers of the National Institutes of Health, who recommended additional concepts and measures for consideration.

Using these three frameworks and recommendations, the Task Force adopted the following as the guiding definition for the survey:

“Social determinants of health are the conditions and context in which people are born, live, learn, play, work, and worship across the lifespan that influence quality of life.”

### Cognitive and online pilot testing

Candidate measures that were considered for inclusion in the survey underwent cognitive interviewing and pilot testing prior to finalizing final instruments and items.

Prior to launching the SDOH survey, a pilot study was conducted from January 2020 - February 2020 and February 2021 - April 2021. The methods of the pilot study included cognitive interviews and online testing, similar to the methods described in the development of the baseline surveys for *All of Us*.^15^ The pilot study included qualitative and quantitative assessment of the SDOH survey in English and Spanish versions, the two languages that are currently used in *All of Us*. The pilot study recruited individuals aged 18 and older with a focus on ensuring the sample included the perspectives of UBR populations, particularly those underrepresented due to racial and ethnic identity, income, and preferred language. The Pilot Research Core ‘s Expression of Interest Registry, Latino Center of the Midlands in Omaha, Nebraska, and Cint (https://www.cint.com/), an online survey audience platform, served as the sole recruitment methods for reaching key populations to test the survey.

### Survey modifications after testing

The cognitive interviews provided feedback on the content and the processes for fielding the survey, which was incorporated in the final SDOH survey (Appendix 2). Content modifications that were implemented included eliminating some concepts from the survey. Process recommendations for fielding the survey that were implemented included providing materials so that participants who indicated social needs via survey could be made aware of relevant resources for the issues they reported. Online pilot testing of the survey demonstrated completion times that were in the range of 10-15 minutes. Pilot completion times were similar among different demographic or language groups (Spanish or English).

### Survey approval and release

The final SDOH survey was IRB-approved June 23, 2021 and launched on November 1, 2021. The SDOH concepts approved in the final survey are listed in Table 1. The SDOH survey is optional and available in English and Spanish language versions to all participants who have completed the first three *All of Us* surveys (The Basics, Overall Health, and Lifestyle). Participants select the language in which they take the survey at the time of program enrollment.

**Table 1.**
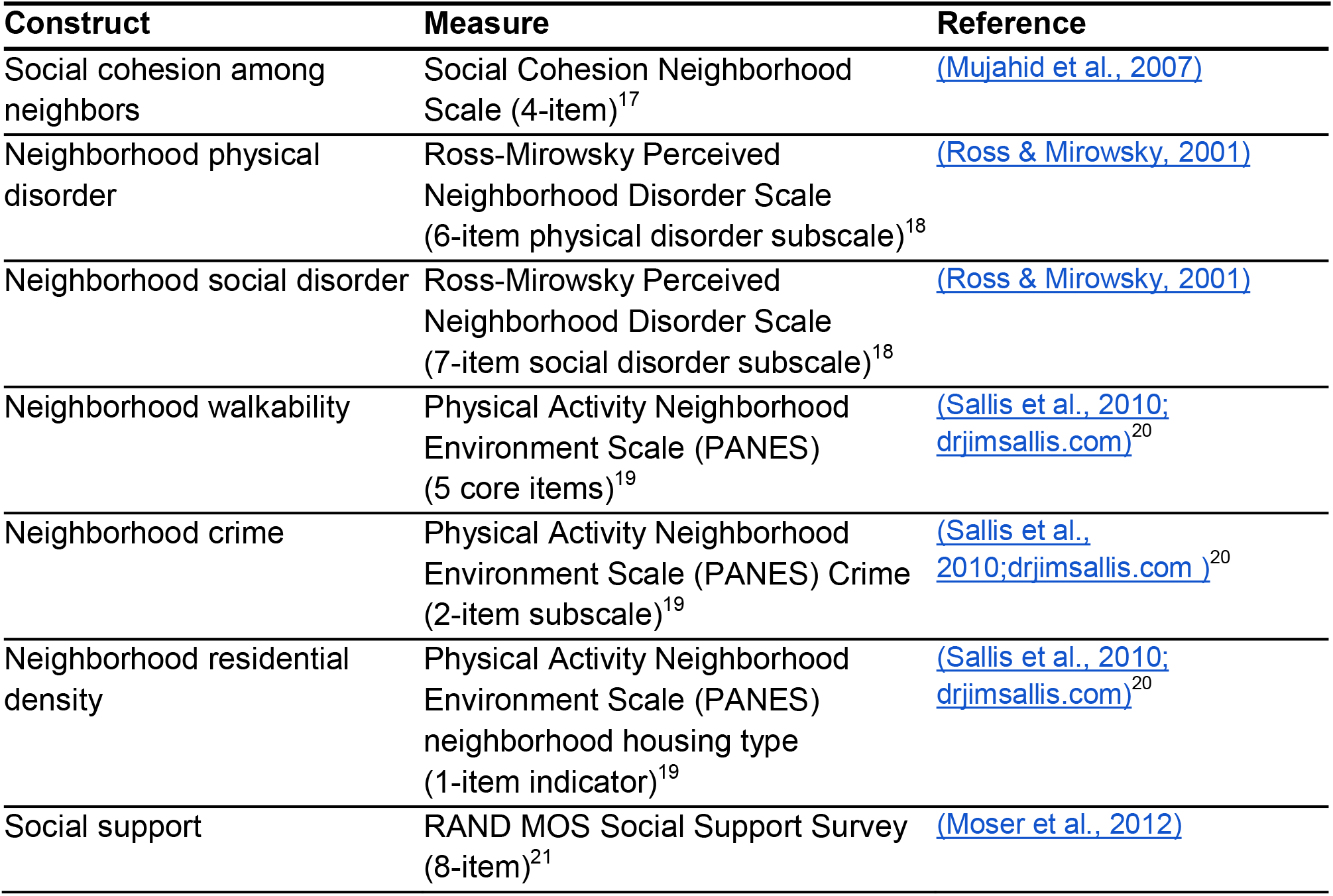
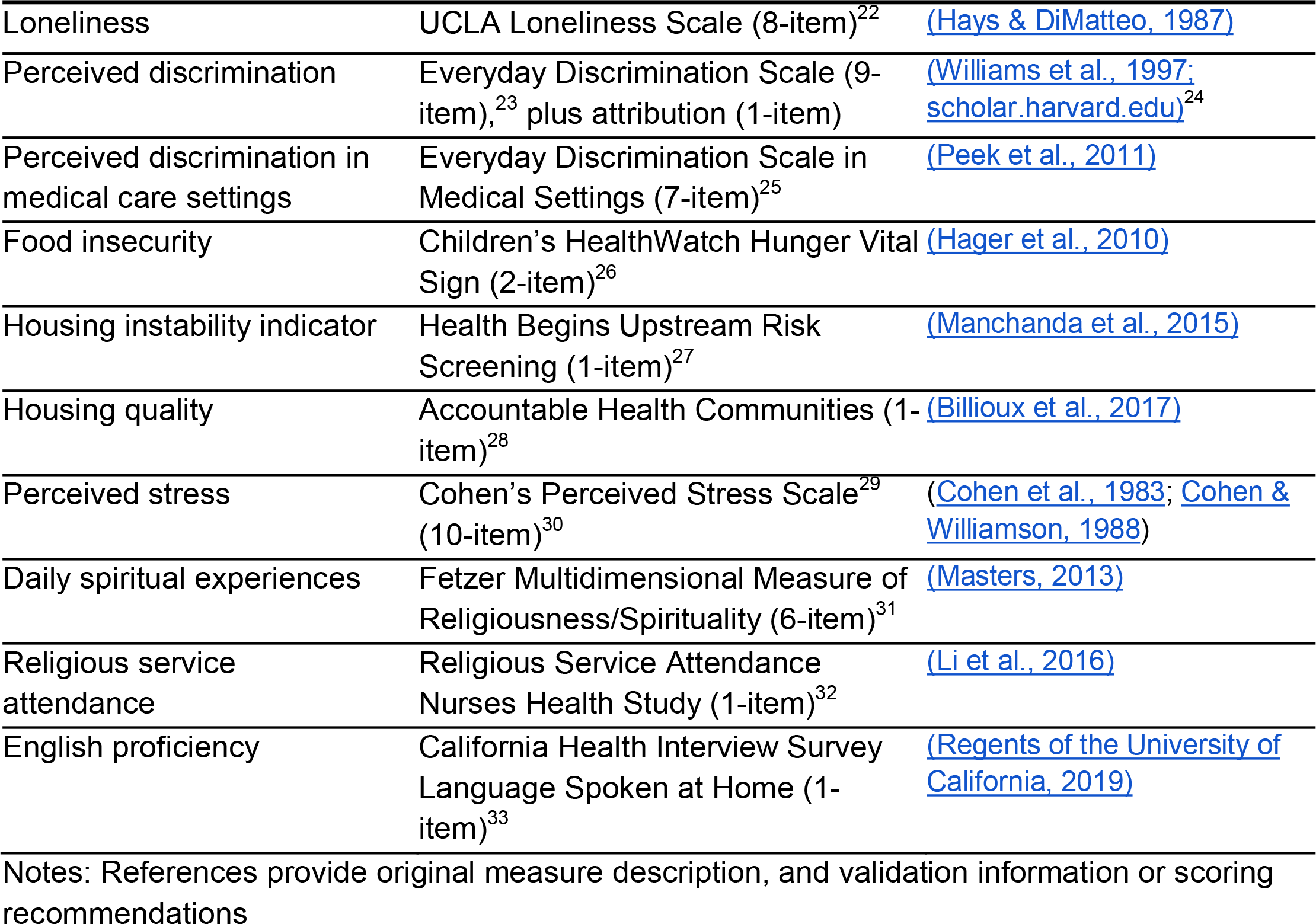
Constructs and Measures in the *All of Us* SDOH survey.

### Accessing data from the SDOH survey

The SDOH survey is publicly available online.^16^ The data can be found on the on-line workbench at the following link: http://www.researchallofus.org/data-tools/workbench/. The SDOH survey questions, responses with answer concept ids, number, and percentages of participants who selected each response, along with bar charts showing the number of participants who chose each answer by the sex assigned at birth and age when the survey was taken, are publicly available via an on-line Data Browser at: (https://databrowser.researchallofus.org/).

### Construction of scales

We constructed scales and scored items in alignment with validated usage in the literature. Two exceptions include the presentation of Religious Service Attendance, and Daily Spiritual Experiences scales, which incorporated *All of Us* Participant Ambassador feedback by adding response set options for participants to indicate “I am not religious,” or to indicate “a higher power,” as an alternative conceptualization of spiritual experience. Due to a transcription error for the Religious Attendance measure, an incorrect response set was displayed for 11,795 participants; instructions to identify these observations are described in Appendix 3. A detailed scoring recommendation for all measures is included in Appendix 3.

### Statistical analyses

Characteristics of *All of Us* participants who responded to the SDOH survey, and those who were eligible but did not submit responses to the SDOH survey, were summarized in means and standard deviations (SD) for continuous variables, and counts and percentages for categorical variables. Among those who responded to the SDOH survey, distributions of the constructed scales were reported in mean, SD, range, median, and interquartile range (IQR). Item non-response was examined by demographic category to understand potential bias in missing data. Item non-response was defined as insufficient items completed by a participant to calculate a scale. In most cases, a scale could not be calculated with more than one or two items not completed; item non-response definitions for each scale are provided in Appendix 3. For each scale, Cronbach ‘s alpha coefficients were used to examine internal consistency overall and by demographic category. Categories with less than 20 participants were masked to preserve privacy.

Multivariable logistic regression models were used to examine if demographic categories were associated with item non-response. The predictors in the model included racial identity, sex assigned at birth, gender identity, sexual orientation, educational attainment, annual household income, disability status, and current age. Age was modeled with a restricted cubic spline function with three knots. Each model is displayed visually as a forest plot to compare the odds ratios and corresponding confidence intervals (CIs). For each plot, the odds of an individual of a particular demographic category (racial identity, sexual orientation, etc.) not responding to the survey scale of interest are compared to the odds of non-response for an indicated reference population within that variable; this yields an odds ratio of item non-response comparing the two groups. Since age was treated non-linearly, two odds ratios were calculated for age: people aged 25 and 75 were compared to a reference group of age 50 for illustration purposes.

The ‘pandas’, and ‘numpy’ Python packages were used for data cleaning, and the ‘pingouin’ package was used to calculate Cronbach’s alpha for the SDOH scales. The R kernel within the Jupyter environment was used for the remaining analysis. In particular, the 95% CIs for proportion differences were constructed using *prop*.*test* function in the ‘stats’ (version 4.2.0) package, regression models were performed using the ‘rms’ (version 6.6) package, and forest plots were generated by the ‘metafor’ (version 4.0) packages.

The current analyses are conducted as part of a demonstration project designed to describe *All of Us* cohort data in preparation for releasing data to the *All of Us* Researcher Workbench. This demonstration project began work in the version 6 data. The data described in this report are results from the *All of Us* Researcher Workbench version 7 released to the Researcher Workbench in April 2023. The work described here was proposed by Consortium members and confirmed as meeting criteria for non-human subjects research by the *All of Us* Institutional Review Board. Results reported are in compliance with the *All of Us* Data and Statistics Dissemination Policy disallowing disclosure of group counts under 20.

## Results

### SDOH survey questions

The constructs and measures in the final fielded SDOH survey are listed in Table 1.

### Description of survey participants

A total of 397,732 participants were eligible to complete the SDOH survey, of which 332,986 (83.7%) were UBR. As of June 30, 2022 a total of 117,783 (29.6%) participants provided any SDOH survey data, of which 92,300 (78.4%) were UBR.

There were differences in the characteristics of those who answered any portion of the survey (survey respondents) and those who did not answer any portion of the survey (non-respondents, Table 2). Survey respondents were predominantly White as compared to non-respondents (74.6% vs 44.8%). More respondents had a college or advanced degree than non-respondents (62.5% vs. 36.0%). Survey non-respondents had lower incomes ($50,000 or less per year) compared to survey respondents (45.0% vs. 27.8%). Intersex, none of these describe me, prefer not to answer, or skip Gender identity 2,978 (2.5%) 5,384 (1.9%

**Table 2:**
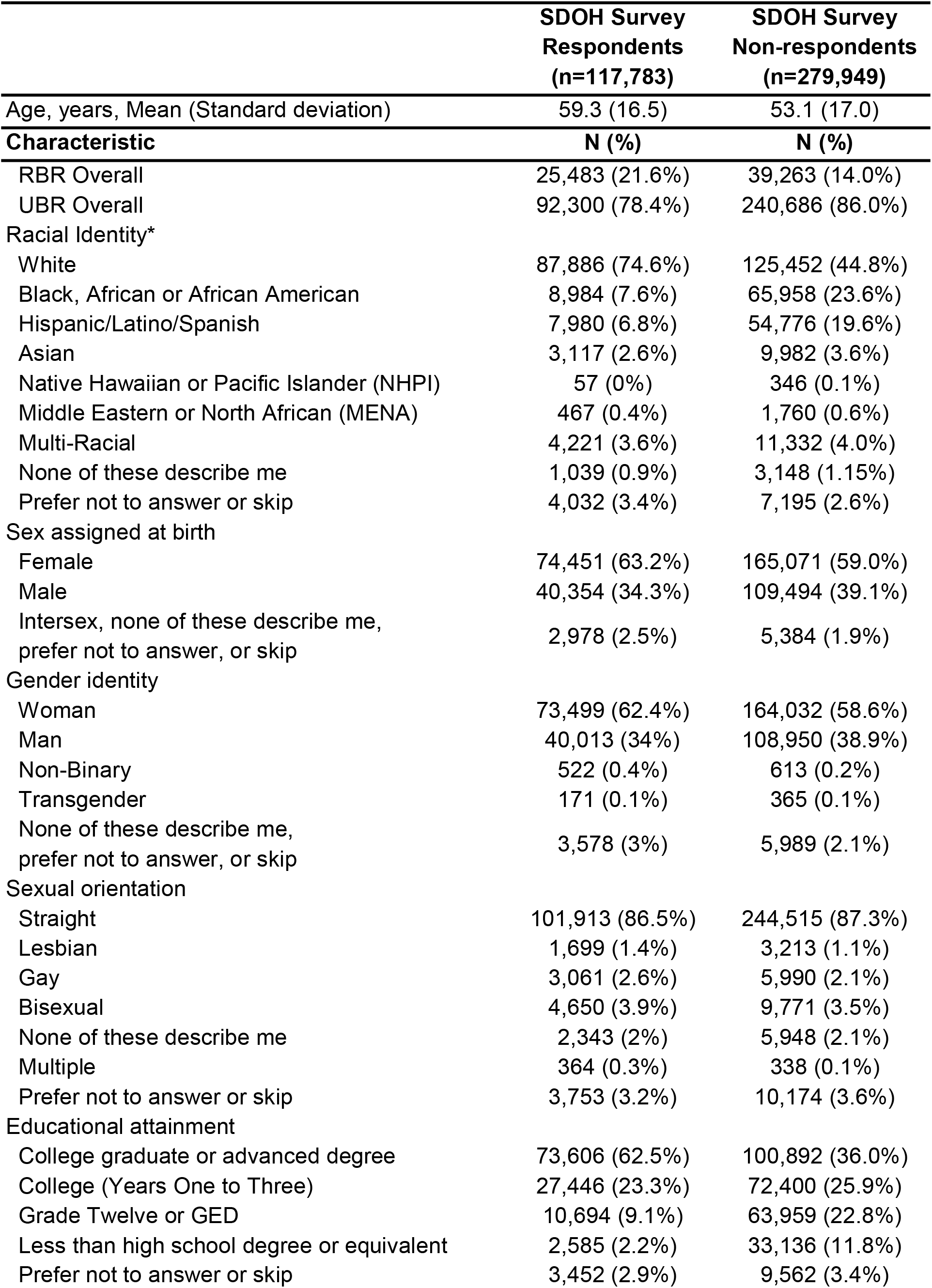
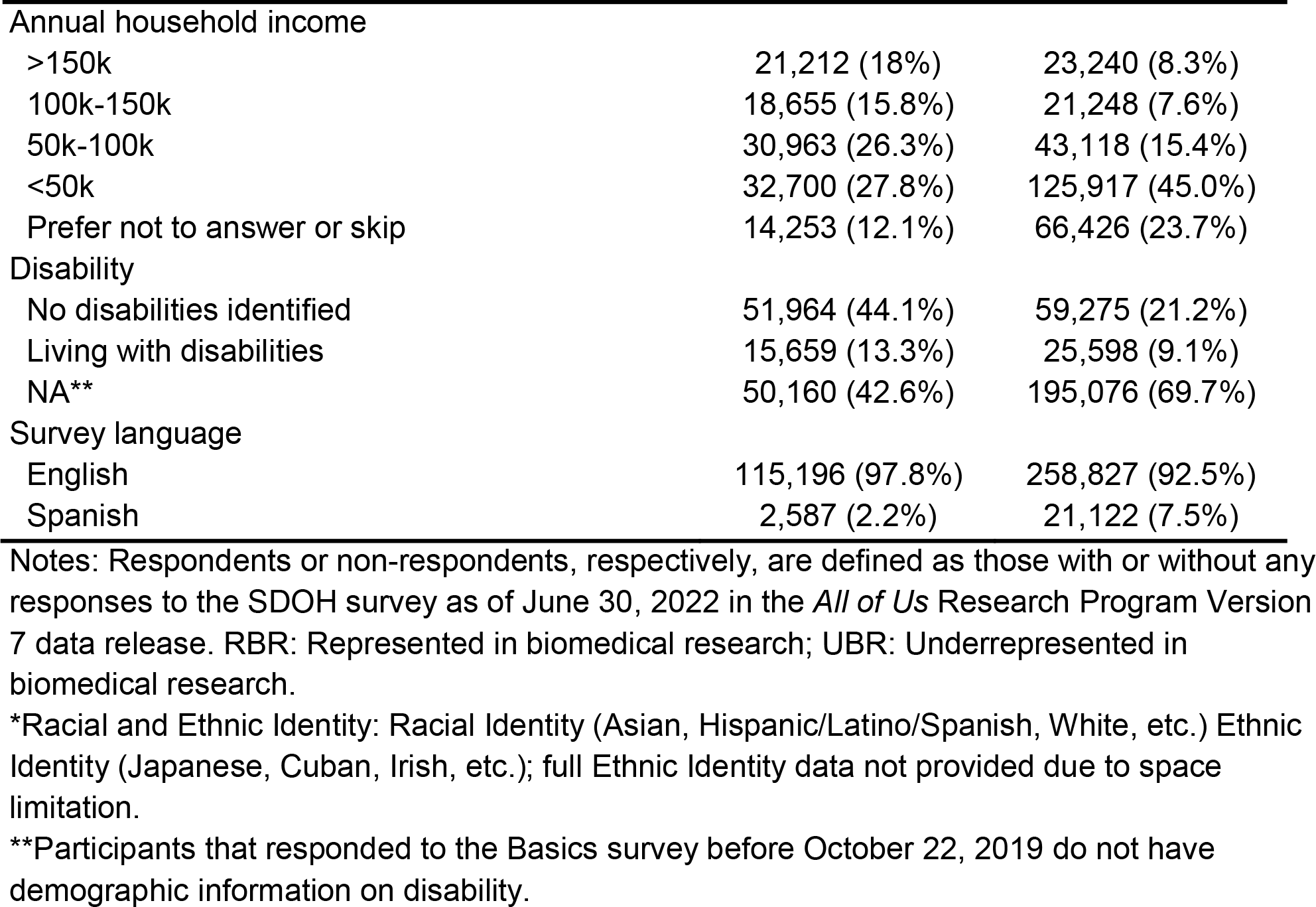
Demographic characteristics of participants.

|  | **SDOH Survey**<br>**Respondents**<br>**(n=117,783)** | **SDOH Survey**<br>**Non-respondents**<br>**(n=279,949)** |
| --- | --- | --- |
| Age, years, Mean (Standard deviation) | 59.3 (16.5) | 53.1 (17.0) |
| **Characteristic** | **N (%)** | **N (%)** |
| RBR Overall | 25,483 (21.6%) | 39,263 (14.0%) |
| UBR Overall | 92,300 (78.4%) | 240,686 (86.0%) |
| Racial Identity* |  |  |
| White | 87,886 (74.6%) | 125,452 (44.8%) |
| Black, African or African American | 8,984 (7.6%) | 65,958 (23.6%) |
| Hispanic/Latino/Spanish | 7,980 (6.8%) | 54,776 (19.6%) |
| Asian | 3,117 (2.6%) | 9,982 (3.6%) |
| Native Hawaiian or Pacific Islander (NHPI) | 57 (0%) | 346 (0.1%) |
| Middle Eastern or North African (MENA) | 467 (0.4%) | 1,760 (0.6%) |
| Multi-Racial | 4,221 (3.6%) | 11,332 (4.0%) |
| None of these describe me | 1,039 (0.9%) | 3,148 (1.15%) |
| Prefer not to answer or skip | 4,032 (3.4%) | 7,195 (2.6%) |
| Sex assigned at birth |  |  |
| Female | 74,451 (63.2%) | 165,071 (59.0%) |
| Male | 40,354 (34.3%) | 109,494 (39.1%) |
| Intersex, none of these describe me,<br>prefer not to answer, or skip | 2,978 (2.5%) | 5,384 (1.9%) |
| Gender identity |  |  |
| Woman | 73,499 (62.4%) | 164,032 (58.6%) |
| Man | 40,013 (34%) | 108,950 (38.9%) |
| Non-Binary | 522 (0.4%) | 613 (0.2%) |
| Transgender | 171 (0.1%) | 365 (0.1%) |
| None of these describe me,<br>prefer not to answer, or skip | 3,578 (3%) | 5,989 (2.1%) |
| Sexual orientation |  |  |
| Straight | 101,913 (86.5%) | 244,515 (87.3%) |
| Lesbian | 1,699 (1.4%) | 3,213 (1.1%) |
| Gay | 3,061 (2.6%) | 5,990 (2.1%) |
| Bisexual | 4,650 (3.9%) | 9,771 (3.5%) |
| None of these describe me | 2,343 (2%) | 5,948 (2.1%) |
| Multiple | 364 (0.3%) | 338 (0.1%) |
| Prefer not to answer or skip | 3,753 (3.2%) | 10,174 (3.6%) |
| Educational attainment |  |  |
| College graduate or advanced degree | 73,606 (62.5%) | 100,892 (36.0%) |
| College (Years One to Three) | 27,446 (23.3%) | 72,400 (25.9%) |
| Grade Twelve or GED | 10,694 (9.1%) | 63,959 (22.8%) |
| Less than high school degree or equivalent | 2,585 (2.2%) | 33,136 (11.8%) |
| Prefer not to answer or skip | 3,452 (2.9%) | 9,562 (3.4%) |

**Table 2: Demographic characteristics of participants**
|  | **SDOH Survey**<br>**Respondents**<br>**(n=117,783)** | **SDOH Survey**<br>**Non-respondents**<br>**(n=279,949)** |
| --- | --- | --- |
| Annual household income |  |  |
| >150k | 21,212 (18%) | 23,240 (8.3%) |
| 100k-150k | 18,655 (15.8%) | 21,248 (7.6%) |
| 50k-100k | 30,963 (26.3%) | 43,118 (15.4%) |
| <50k | 32,700 (27.8%) | 125,917 (45.0%) |
| Prefer not to answer or skip | 14,253 (12.1%) | 66,426 (23.7%) |
| Disability |  |  |
| No disabilities identified | 51,964 (44.1%) | 59,275 (21.2%) |
| Living with disabilities | 15,659 (13.3%) | 25,598 (9.1%) |
| NA** | 50,160 (42.6%) | 195,076 (69.7%) |
| Survey language |  |  |
| English | 115,196 (97.8%) | 258,827 (92.5%) |
| Spanish | 2,587 (2.2%) | 21,122 (7.5%) |

Notes: Respondents or non-respondents, respectively, are defined as those with or without any responses to the SDOH survey as of June 30, 2022 in the *All of Us* Research Program Version 7 data release. RBR: Represented in biomedical research; UBR: Underrepresented in biomedical research.
\*Racial and Ethnic Identity: Racial Identity (Asian, Hispanic/Latino/Spanish, White, etc.) Ethnic Identity (Japanese, Cuban, Irish, etc.); full Ethnic Identity data not provided due to space limitation.
\*\*Participants that responded to the Basics survey before October 22, 2019 do not have demographic information on disability.

### Internal consistency reliability of scales

Internal consistency measured by Cronbach’s alpha was over 0.8 for almost all scales (Table 3). The multi-dimensional Physical Activity and Neighborhood Environment (PANES) Walking and Bicycling scale had the lowest Cronbach’s alpha at 0.78. Internal consistency did not vary substantially by participant characteristics for most scales (Appendix 3). However, for participants who identify as Native Hawaiian and Pacific Islander (for whom the sample size was N = 57), the PANES Walking and Bicycling scale had a much lower Cronbach’s alpha (0.58) compared to that of other groups where alpha coefficients for the Walking and Bicycling scale ranged from 0.70 to 0.79. Estimates of internal consistency on other scales for Native Hawaiian Pacific Islanders ranged from 0.78 to 0.95.

**Table 3:**
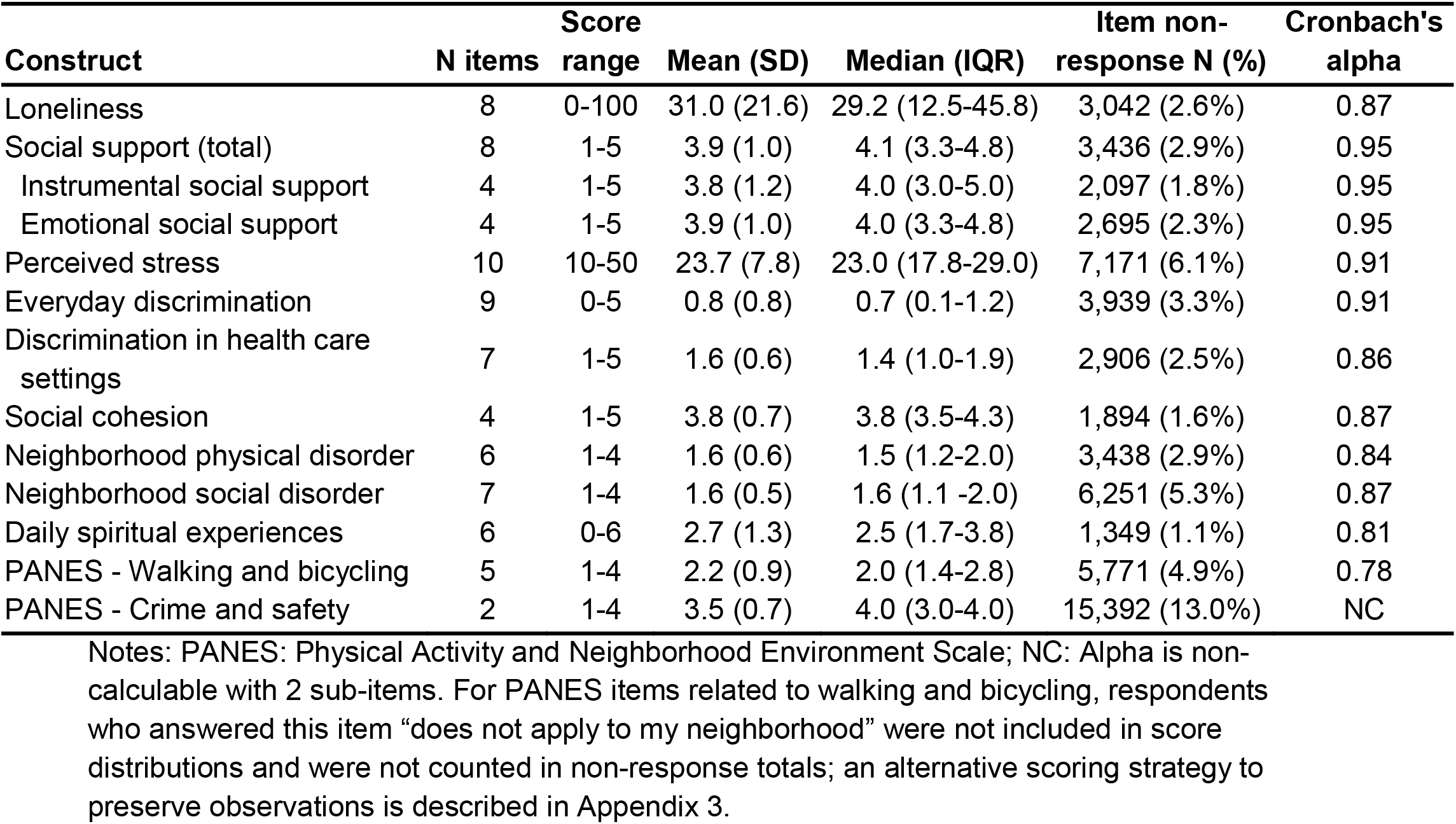
Score distributions, item non-response and Cronbach’s alpha (N=117,783)

### Item non-response and scale score distributions among survey respondents

Item non-response was infrequent. The majority of SDOH survey scales had fewer than 5% missing data due to item non-response (Table 3). The PANES Crime and Safety scale had the highest proportion of item non-response at 13.0%. The Cohen Perceived Stress Scale, the Neighborhood Social Disorder Scale, and the single housing quality item also had greater than 5% item non-response at 6.1%, 5.3%, and 5.2% respectively. Score distributions for scales are described in Table 3. Item non-response for the single Religious Service Attendance item was 1.5% (N=1,825). A single-item indicator from the PANES instrument for neighborhood residential density/housing type had 2.1% item non-response (N=2,475). Item non-response among the categorical measures was 1.3% (N= 1,542) for food insecurity, 3.1% (N = 3,620) for housing instability, and 5.2% for housing quality problems (N = 6,134). Of the respondents for the categorical items, 13.5% had food insecurity, 2.7% had housing instability assessed as multiple address changes in 12 months, and 21.1% had housing quality problems such as bug infiltration, mold, or lead pipes.

Item non-response varied most by educational attainment (Figure 1), racial identity (Figure 2), and survey language (Figure 2). For example, in the Loneliness scale, Black, African, and African American participants and Hispanic/Latino/Spanish participants had 5.1% and 5.7% item-non response respectively compared to 2.0% item-non response among White participants. Participants with less than a high school degree or equivalent had 8.6% item non-response for the Loneliness scale compared to 1.9% among participants with a college or advanced degree. Participants who took the SDOH survey in Spanish had 10.6% item non-response for the Loneliness scale compared to 2.4% among those who took it in English. In the PANES Crime and Safety scale, which had higher item non-response than other scales, Black, African, and African American participants and Hispanic/Latino/Spanish participants had % and 17.1% item non-response respectively compared to 12.1% among White participants. Participants with less than a high school degree or equivalent had 21.3% item non-response compared to 11.8% among participants with a college or advanced degree. Participants who took the PANES Crime and Safety scale in Spanish had 25.6% item non-response compared to 12.7% item non-response among those who took it in English. A description of absolute differences in item non-response for each scale by participant characteristics is provided in Appendix 4.

**Figure 1:**
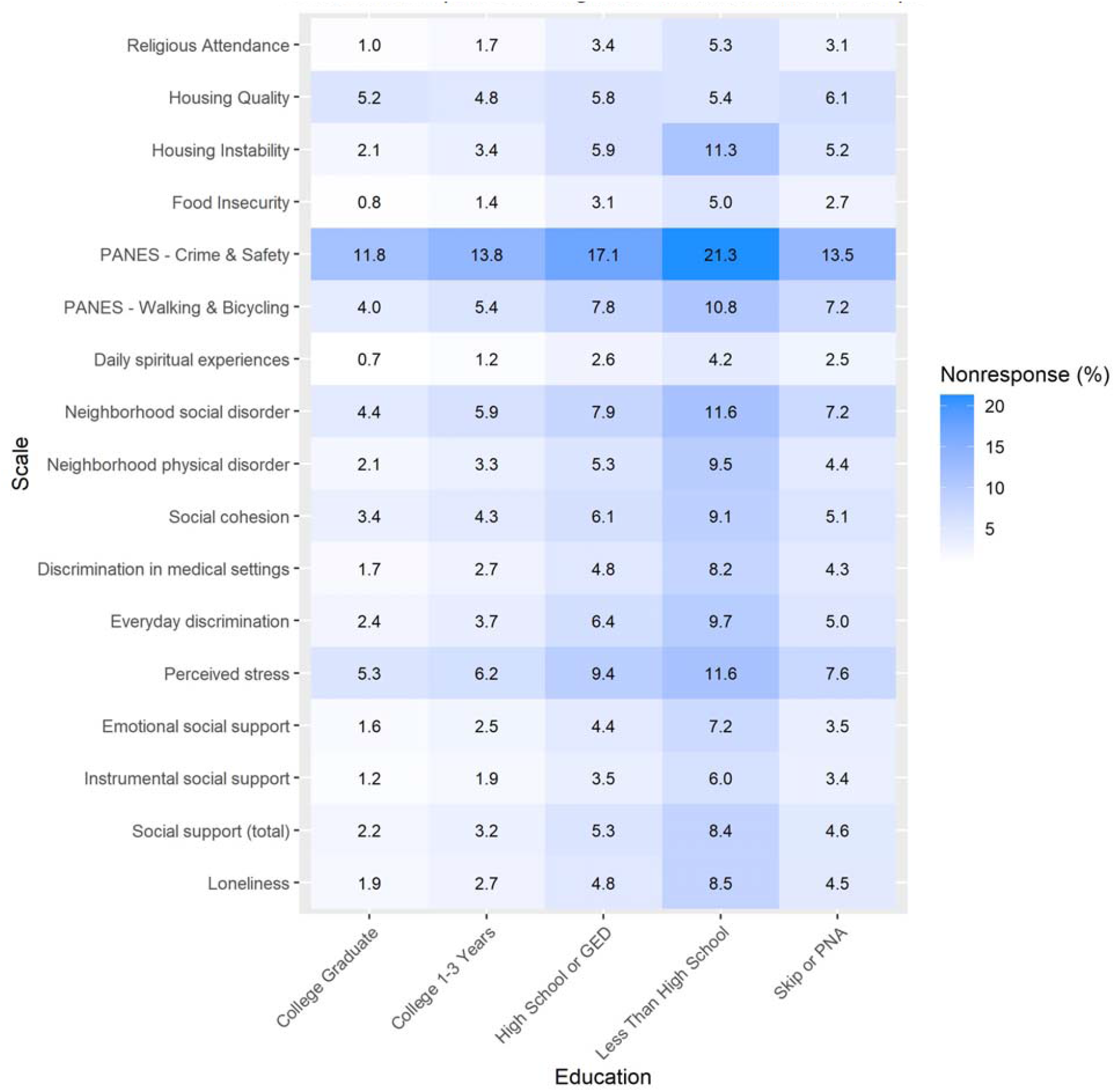
Item non-response percentage of the SDOH scales by educational attainment. Notes: Data represent the percentage of SDOH survey respondents with an incalculable scale due to item non-response, by educational attainment. PANES: Physical Activity and Neighborhood Environment Scale; PNA: Prefer not to answer. Participants who responded to the incorrect response set for the Religious Service Attendance item (N=11,795) are flagged as ‘invalid’ in version 7 data; these respondents are not included in item non-response calculations.

**Figure 2:**
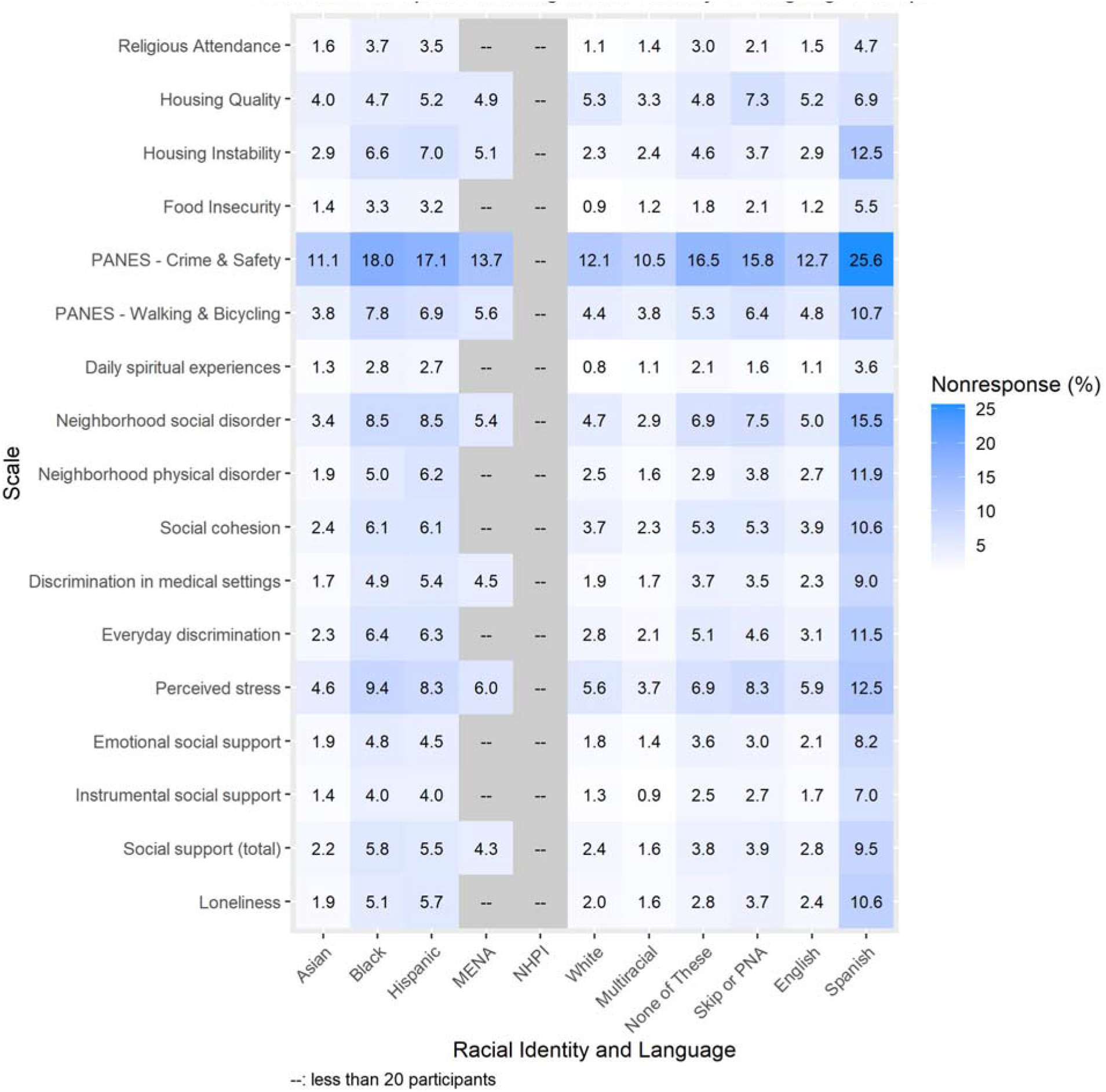
Item non-response percentage of the SDOH scales by Racial Identity and Language Groups (English and Spanish). Notes: Data represent the percentage of SDOH survey respondents with an incalculable scale due to item non-response, by Racial Identity and Language group. Participants who responded to the incorrect response set for the Religious Service Attendance item (N=11,795) are flagged as ‘invalid’ in version 7 data; these respondents are not included in item non-response calculations. PANES: Physical Activity and Neighborhood Environment Scale; Hispanic: Hispanic/Latino/Spanish MENA: Middle Eastern or North African; NHPI: Native Hawaiian or Pacific Islander; PNA: Prefer not to answer Data shaded in gray are suppressed due to small sample size < 20

### Multivariable logistic regression models

The odds of item non-response for any scale are shown in Figure 3. Black, African, or African-American (OR=1.55, 95% CI:1.48-1.63) and Hispanic/Latino/Spanish (OR=1.59, 95% CI:1.51-1.68) participants had higher item non-response compared to White participants. Item non-response was higher amongst those with lower educational attainment compared to those with a college or advanced degree. Item non-response was also lower for those with higher income levels compared to those making less than $50,000 per year. Compared to an individual aged 50, participants aged 25 had less item non-response (OR=0.56, 95% CI:0.54-0.59) and participants aged 75 had greater item non-response (OR=2.60, 95% CI:2.53-2.67). Bisexual (OR=0.86, 95% CI: 0.80-0.93) participants had lower odds of item non-response for any scale than straight participants.

**Figure 3:**
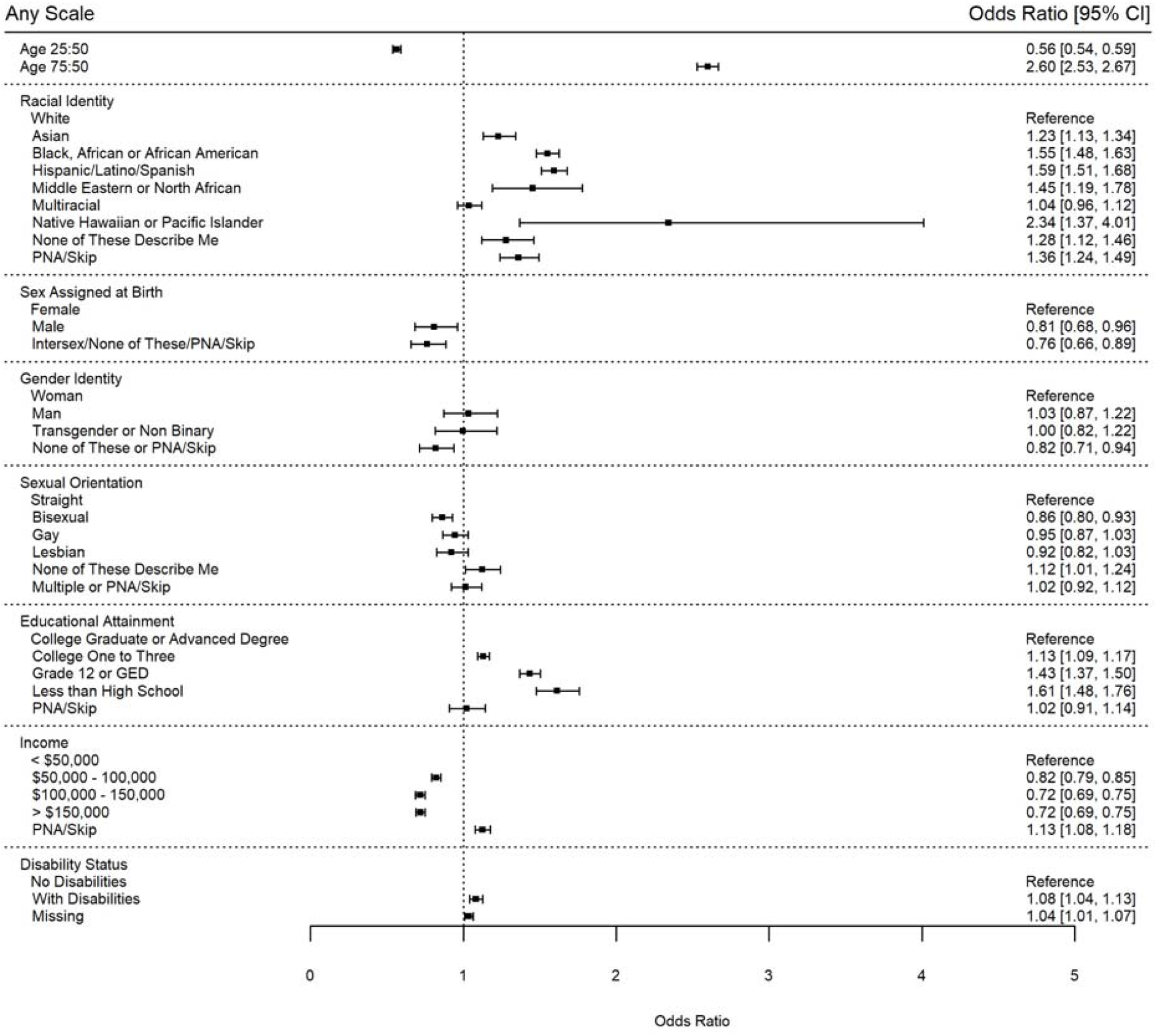
Odds of item non-response or incalculable score for any of the seventeen SDOH scales. Notes: Black: Black, African or African-American; Hispanic: Hispanic/Latino/Spanish; PNA: Prefer not to answer

Several patterns stood out across all models (Appendix 5). For most scales, patterns in item non-response were seen by racial identity, educational attainment, income level, and age. As an example, in the Perceived Stress scale, Black, African or African-American participants (OR=1.78, 95% CI: 1.64-1.93) and Hispanic/Latino/Spanish participants (OR=1.70, 95% CI: 1.55-1.87) had a higher odds of item non-response compared to White participants; participants with less than a high school education were 1.79 (95% CI: 1.55-2.06) times more likely to have item non-response compared to participants with a college or advanced degree; participants with an income level of over $150,000 per year were 0.73 (95% CI: 0.67-0.80) times as likely to have item non-response compared to participants with an income level of less than $50,000 per year; and participants aged 25 years old were 0.60 (95% CI: 0.54-0.66) times as likely and participants aged 75 years old were 2.71 (95% CI: 2.59-2.84) times more likely to have item non-response compared to an individual aged 50 years old.

The food insecurity item, housing instability item, PANES Crime and Safety items, and housing quality item had atypical patterns in item non-response compared to other scales. For food insecurity, both a typical 25-year old participant (OR=1.34, 95% CI: 1.15-1.57) and a typical 75-year old (OR=1.56, 95% CI: 1.41-1.73) participant had higher odds of item non-response when compared to a 50-year old participant. In housing instability, all racial identities other than the “prefer not to answer or skip” group had significantly higher odds of item non-response than White participants. Transgender and non-binary participants had lower odds of item non-response (OR=0.40, 95% CI 0.17-0.93) than those who identified as a woman. Both bisexual (OR=0.74, 95% CI 0.58-0.95) and gay (OR=0.66, 95% CI 0.50-0.85) participants had lower odds of item non-response than straight participants. However, those who did not answer the sexual orientation question had higher odds of item non-response missingness (OR=1.33, 95% CI 1.09-1.63) than straight respondents. For the PANES Crime and Safety items, all UBR racial identity groups had higher odds of item non-response than White participants. Typically, participants living with at least one identified disability had slightly higher odds of item non-response than those without identified disabilities, but in the housing quality item, participants living with a disability had lower odds of item non-response than than those without identified disabilities (OR=0.86, 95% CI 0.79-0.94).

## Discussion

The SDOH survey demonstrated good to excellent internal consistency in measurement of several SDOH concepts and within multiple diverse population groups, including those who are underrepresented in biomedical research. Among those who attempted the survey, data collection was fairly complete with low item non-response (0.9% to 13.5% missing) for most scales. Item non-response varied by SDOH measure and by demographic categories, notably, with differences in item non-response by racial identity, educational attainment, and survey language. Taken together, the relatively complete data among survey respondents and the internal consistency of scales are promising for the use of the survey to better understand the role of the measured SDOHs in precision medicine research. Importantly, patterns of survey non-response and missingness are documented in this report to assist the researcher with managing bias due to differential survey response patterns in the current version 7 release. We note that few participants (2.2%) completed the survey in Spanish, and additional data are needed to confirm reliability and generalizability for administering the survey in Spanish.

The SDOH survey has limitations. The survey was designed to elicit participants’ perceptions of their social environments, psychosocial connections and conditions of their daily lives that would be applicable to multiple research studies and across several complex models of disease or health promotion. The survey does not measure or explore *structural social determinants of health inequities* that detail “the mechanisms through which social hierarchies and social conditions are created”.^1^ For example, the survey sought to provide measures of perceived discrimination, but was not designed to gather hypothesis-driven information from participants on structural racism or policies that lead to participants’ experiences of unequal treatment. To facilitate further assessment of structural social determinants of health, the *All of Us* Research program will conduct geocoding, and plan future assessment of area-level and other contextual measures to provide insight into macro-level social conditions of relevance to precision medicine. Additionally, the expansive scope of social experiences that participants may perceive as important to the condition of their lives is considerable, and all relevant concepts could not feasibly be included in one survey. For example, the Task Force deferred capturing more detailed measures of wealth and acculturation, for potential future assessment in dedicated data collection efforts. We note that other *All of Us* surveys currently measure income, home ownership, education, employment status, health literacy, and other topics related to SDOH, which are also available in the version 7 release. Continued engagement with *All of Us* participants and scientists is warranted to develop the next set of surveys that deepen scientific understanding of social concepts that influence health.

While data on internal consistency and data completeness are presented in this report, additional tests of psychometric properties of scales in the *All of Us* cohort should continue to be assessed in hypothesis-driven research. Importantly, the SDOH survey remains in the field, and as of April 2023, 41% of those eligible have now completed this survey. As the survey is fielded more completely, metrics on survey non-response and updates on survey performance must continue to be monitored and evaluated to gauge internal reliability and external generalizability of SDOH data.

## Summary

The *All of Us* SDOH survey, developed through engagement with scientists and *All of Us* participant partners, has items and scales with strong psychometric properties that measure social experiences among a large, diverse participant population, for the purpose of advancing precision medicine research. Additional work should investigate the construct validity of some social concepts by geography, and within specific groups, including Spanish language users, and in Native Hawaiian and Pacific Islander groups, where scale internal consistency reliability may have varied compared to other groups with larger sample sizes. Future surveys should add to the breadth of concepts that can be explored in *All of Us*, and where available, researchers should compare *All of Us* data with other cohorts to enhance our understanding of social experiences relevant to precision medicine research.

## Data Availability

The SDOH survey is publicly available online. The data can be found on the on-line workbench at the following link: https://www.researchallofus.org/data-tools/workbench/

https://databrowser.researchallofus.org/

## Acknowledgements

We thank our colleagues, Brandy Mapes, Ashley Green and Chris Lord for providing their support and input throughout the demonstration project lifecycle. We thank Jun Qian for providing input on the project’s code review. We thank the DRC’s Research Support team for their help during implementation. We also thank the *All of Us* Science Committee and *All of Us* Steering Committee for their efforts evaluating and finalizing the approved demonstration projects. The *All of Us* Research Program would not be possible without the partnership of contributions made by its participants. See Appendix 6 for a roster of past and present *All of Us* principal investigators. To learn more about the *All of Us* Research Program’s research data repository, please visit https://www.researchallofus.org/.

The *All of Us* Research Program is supported by the National Institutes of Health, Office of the Director: Regional Medical Centers: 1 OT2 OD026549; 1 OT2 OD026554; 1 OT2 OD026557; 1 OT2 OD026556; 1 OT2 OD026550; 1 OT2 OD 026552; 1 OT2 OD026548; 1 OT2 OD026551; 1 OT2 OD026555; IAA#: AOD 16037; Federally Qualified Health Centers: 75N98019F01202.; Data and Research Center: 1 OT2 OD35404; Biobank: 1 U24 OD023121; The Participant Center: U24 OD023176; Participant Technology Systems Center: 1 OT2 OD030043; Community Partners: 1 OT2 OD025277; 3 OT2 OD025315; 1 OT2 OD025337; 1 OT2 OD025276. In addition, the *All of Us* Research Program would not be possible without the partnership of its participants.

