## Appendix 1 for "Measuring Social Determinants of Health in the *All of Us* Research Program: Technical Document"

### Appendix Table 1: SDOH Survey Development Process

| Process | Considerations |
| --- | --- |
| Phase I: Select conceptual frameworks and define SDOH for the *All of Us* survey | - Prioritize frameworks that guide research on connections between social factors and health - Prioritize frameworks and definitions that improve communication on social concepts in large and diverse participant audiences |
| Phase II: Define inclusion and exclusion criteria, and priorities for selecting and using constructs and measures | Inclusion criteria.  Concepts should:   - measure *perceptions* that can only be collected through participant responses; - connect to core drivers of health inequities (e.g., perceived discrimination); - have documentation on measure validation and psychometric performance; - have strong use cases (structural, social and biologic) to facilitate research on mechanisms between SDOHs and health.   Exclusion criteria.  Concepts should not be:   - new concepts and measures without psychometric validation (with rare exception); - concepts that can be collected without burdening participants (e.g., via geocoding) - concepts that may be more reliability captured through other modalities besides participant surveys (e.g., wealth) - concepts that may require sufficient items or measures to merit a dedicated survey (e.g., acculturation, wealth)   Priorities for selecting and incorporating measures.   - constructs and measures should be validated with high reliability in diverse cohorts and in multiple languages. - measures should be included in the form in which they were validated, and item response sets should not be altered as practically possible. |
| Phase III: Review the science to select standardized measures with use cases in precision medicine | - Prioritize measures that operationalize concepts in the World Health Organization Conceptual Framework for Action on the Social Determinants of Health,^1^ and the five domain areas of the Healthy People framework for SDOH (*social and community*; *economic stability*; *education*; *neighborhood and built environment*; *health and health care*.)^12^ - Prioritize measures with data from epidemiologic cohort studies and other designs that elucidate mechanisms among SDOHs and connections to health |
| Phase IV: Examine surveys, measures and items in other large biobanks, cohort studies, epidemiologic surveys and toolkits to find opportunities to align measures | - UK Biobank - Million Veteran Program - Behavioral Risk Factor Surveillance Survey - National Health and Nutrition Examination Survey - [NIH PhenX Toolkit SDOH Collections](https://www.phenxtoolkit.org/collections/sdoh) |
| Phase V: Coordinate internally with other *All of Us* task forces to avoid duplication | - Mental health - Environmental health - “The Basics” assessment of income, educational attainment, race, ethnicity, age, sexual orientation, gender identity, health care access, insurance status |
| Phase VI: Consult with scientific subject matter experts and participant partners in *All of Us* | - Developers of key measures of interest - NIH Institutes: NHLBI, NIDDK, NICHD, NIMHD, OBSSR, ORWH - *All of Us* Participant Ambassadors |
