## Appendix 2 for "Measuring Social Determinants of Health in the *All of Us* Research Program: Technical Document"

### Appendix Table 2: SDOH Survey Modifications

Survey modifications based on cognitive interview and *All of Us* Participant Ambassador feedback

Prioritized measures for inclusion and exclusion from the survey

Replaced mentions of “God” with “God (or a higher power)” in measures of religiousness and spirituality.

Added “I am not religious” as a response option in questions pertaining to God.

Added language in introductory text to better define the phrase “social determinants of health”

Re-ordered domains so that questions with negative framing do not all appear at the beginning of the survey.

Added “tool tip” at the beginning of each domain to clarify the scientific value of the survey questions.

Developed participant-facing frequently asked questions (FAQs) to help participants better understand SDOH in general, the topics included in the survey, and the role of SDOH in health and well-being. Some FAQs also include links to additional information for participants who are interested in learning more about SDOH and resources to assist any participants who may be in need of additional information or support in specific SDOH-related areas.
