## Appendix 3 for "Measuring Social Determinants of Health in the *All of Us* Research Program: Technical Document"

### Appendix 3: SDOH Scales and Scoring

SDOH survey scoring documentation and recommendations are provided for the following scales:

**Social Cohesion.** The social cohesion scale connects to theories on neighborhood social organization.^34,35^ The selected scale, also featured in the NIH PhenX toolkit, was modified and validated by Mujahid et al.^17^ There are four items, rated on a 5-point agreement scale (1 = strongly agree, 2 = agree, 3 = neutral (neither agree nor disagree), 4 = disagree, and 5 = strongly disagree). The scoring is the mean of the four items. It was flagged as missing if any of the four items were missing a valid response. The original article reported a Cronbach’s Alpha of 0.74, and a test-retest reliability of 0.65. Validation was done using census tract data.^17^

**Neighborhood Physical and Social Disorder.** The Neighborhood Physical and Social Disorder scale is from the work of Ross and Mirowsky.^18^ The Physical Disorder items reflect negative physical characteristics such as noise, graffiti, and abandoned buildings. The Neighborhood Physical Disorder Scale has six items, with two items reverse scored. Responses are collected using a 4-point agreement scale (1 = strongly disagree; 2 = disagree; 3 = agree; 4 = strongly agree). Scoring is the average of the items, with the score set to missing if more than one item is not a valid response. Neighborhood Social Disorder measures psychosocial problems associated with the people in one’s neighborhood and their behavior (perceived safety, trouble with neighbors). There are seven items in the scale, and it uses a 4-point agreement rating scale (1 = strongly disagree; 2 = disagree; 3 = agree; 4 = strongly agree). The scale is scored by averaging the ratings of the 7 items. The scale is set to missing if more than one item does not have a valid response. Coefficient alpha was 0.92 for the combined physical and social neighborhood disorder measure. The scale was validated against census tract data, health outcomes, and sociodemographic characteristics.^18^

**Physical Activity and Neighborhood Environment Scale (PANES) – Perceived Residential Density/Neighborhood Housing Type**. Sallis et al^19^ developed a series of self-report measure perceptions of the characteristics of the neighborhood environment that can influence physical activity. All of the PANES measures are introduced with the prompt, “Think about the different facilities in and around your neighborhood. By this we mean the area ALL around your home that you could walk to in 10-15 minutes.” Perceived residential density is measured with an indicator variable: “What is the main type of housing in your neighborhood?” There are 5 response choices: “Detached single-family housing; Townhouses, row houses, apartments, or condos of 2-3 stories; Mix of single-family residences and townhouses, row houses, apartments or condos; Apartments or condos of 4-12 stories; and Apartments or condos of more than 12 stories. Interpretation of this item and coding strategies are provided by Sallis et al.^19,36^

**Physical Activity and Neighborhood Environment Scale (PANES) – Walking and Bicycling**. The Walking and Bicycling Scale of the PANES was adapted from Sallis et al^19^ and consists of five items that are rated on a 4-point agreement scale (1 = Strongly disagree; 2 = Somewhat disagree; 3 = Somewhat agree; 4 = Strongly agree; Missing= Skip/Don’t know. Sallis et al^19^ reported adequate test-retest reliability. In this technical report, we scored this scale as the average of the five items and flagged the score as missing if more than one item does not have a valid response. Two items related to walking and bicycling include the response category “Does not apply to my neighborhood.” As an alternative to deleting the “Does not apply to my neighborhood” responses, re-scoring this response category as “1 = Strongly disagree/Does notes apply to my neighborhood” results in good internal consistency reliability in *All of Us* (Cronbach’s alpha =0.78) and may allow the ability to retain observations in rural geographies.

**Physical Activity and Neighborhood Environment Survey (PANES) – Crime Safety.** Two items from the original 17-item PANES scale that represent perceptions of neighborhood safety from crime were selected.^19^ The items are rated on a 4-point agreement scale (1= Strongly disagree; 2 = Somewhat disagree; 3 = Somewhat agree; 4 = Strongly agree; Missing= Skip/Don’t know). Test-retest correlations were reported for individual items and showed acceptable reliability.^19^ We reversed and averaged these items to form a PANES-Crime Safety measure. If either item is not a valid response (including Skip/Don’t know), we flagged the scale as missing. Additional coding strategies for individual items are recommended by Sallis et al.^36^

**Social Support.** The 8-item modified (mMOS-SS) social support scale from the Medical Outcomes Study was used.^21^ The scale measures perceived social support. Each item is rated on a 5-point scale (1 = None of the time; 2 = A little of the time; 3 = Some of the time; 4 = Most of the time; 5 = All of the time). Three scores can be computed using an average of the items within the scale. A total social support scale can be computed using all eight items. It can be computed if 6 or more items have valid responses. The total score has a Cronbach’s Alpha of 0.93. All 8-items had high item-total correlations. There are also two 4-item subscales, one to measure *instrumental support* (e.g., help prepare your meals), and the other to measure *emotional support* (e.g., someone to love and make you feel wanted). Separate reliability coefficients were not reported for the subscales. These scales are computed as averages, and can be used if only a single item is missing, otherwise the scale is set to a missing value. Convergent and discriminant validity was demonstrated using correlations with self-report, demographic, and health measures.^21^

**Loneliness.** The 8-item UCLA Loneliness Scale (ULS-8) was used.^22^ The items measured perceived lack of companionship and social isolation. The Loneliness scale uses a 4-point frequency rating to collect responses (1 = Never; 2 = Rarely; 3 = Sometimes; 4 = Often). The scale is scored by calculating an item mean, and can be computed if 6 or more items have valid responses. Cronbach’s Alpha of 0.84 was reported for the 8-item scale. Convergent validity was evaluated using correlations with other measures of mental health and interpersonal relationships.^22^

**Everyday Discrimination.** A 9-item everyday discrimination scale was developed by Williams et al.^23^ It used a 6-point frequency scale (5 = Almost everyday; 4 = At least once a week; 3 = A few times a month; 2 = A few times a year; 1 = Less than once a year; 0 = Never). Scoring is done by computing the mean item score. The scale is flagged as missing if more than two items do not have a valid score. The coefficient Alpha was 0.88 in the original study. The scale has been validated using self-report health, race, and measures of socioeconomic status.^37^ There is an additional “check all that applies” to a descriptive item that asks participants “What do you think is the main reason for these experiences?” Response options are: 1. Your Ancestry or National Origins, 2. Your Gender, 3. Your Race, 4. Your Age, 5. Your Religion, 6. Your Height, 7. Your Weight, 8. Some other aspect of Your Physical Appearance, 9. Your Sexual orientation, 10. Your education or income level, 11. Other (specify). This item is not scored and is used as descriptive data.

**Discrimination in Health Care Settings.** A 7-item scale to measure experiences of discrimination in health care settings was obtained from Peek et al.^25^ It asks for frequency ratings on a 5-point scale (1 = Never; 2 = Rarely; 3 = Sometimes; 4 = Most of the time; 5 = Always) of how one is treated in health care settings (e.g. You receive poorer service than others). Scoring the scales is done by computing the item average, and the scale is flagged as missing if more than two items do not have valid responses. Cronbach’s Alpha was 0.89, and the test-retest reliability was 0.58. The scale was validated using other measures of discrimination and had low correlations with depression and social desirability.^25^

**Food Insecurity.** A screening Food Insecurity measure based on the work of Hager et al^26^ was included. There are two items (rated as often true, sometimes true or never true). The individual is considered food insecure if any item is rated as sometimes true or often true. High specificity and sensitivity has been reported in classifying families and food insecure using the two-item scale.^38^ Convergent validity was demonstrated using logistic regression models of health outcomes.^26^

**Housing Instability.** A single item assessment of housing instability was included. The item is “In the last 12 months, how many times have you or your family moved from one home to another?” Ash et al^39^ used data on multiple address changes as a measure of housing insecurity/instability to inform payment models for assessing social risks.

**Housing Quality.** An item describing housing instability due to housing quality problems was used in the Accountable Health Communities Screening Tool.^28^ The item is “Think about the place you live. Do you have problems with any of the following?” There are 8 check all that apply response options (1 = Bug infestation; 2 = Mold; 3 = Lead paint or pipes; 4 = Inadequate heat; 5 = Oven or stove not working; 6 = No or not working smoke detectors; 7 = Water leaks; 8 = None of the above). In this report, it is scored as a binary variable and set to zero if option 8 is the only item selected, and to 1 if one or more of options 1-7 are selected. Data on reliability or validity were not available at the time of measure selection.

**Cohen Perceived Stress.** The Cohen Perceived Stress Scale is a 10-item scale^30^ (the original scale had 14 items)^29^ that measures a participant's experience of everyday thoughts and feelings. The questions ask for a frequency rating in the past month using a 5-point scale (1 = Never; 2 = Almost Never; 3 = Sometimes; 4 = Fairly Often; 5 = Very Often). The scale is scored by multiplying the item mean times ten. It can be scored if more than 8 items have a valid response. Cronbach’s Alpha for the original scale was reported to be 0.84 to 0.86.^29^ Validation included associations with life events, depressive symptoms, and healthcare utilization.^29^ The validated 10-item scale had a Cronbach’s Alpha of 0.78.^30^

**Daily Spiritual Experiences.** The Daily Spiritual Experiences scale was adapted from the Brief Multidimensional Measure of Religiousness/Spirituality.^31^ The Daily Spiritual Experiences is a single subscale from a longer instrument and includes items like “I feel God’s presence.” The items are rated using a 6-point scale (6 = Many times a day; 5 = Every day; 4 = Most days; 3 = Some days; 2 = Once in a while; 1 = Never or almost never). In response to *All of* *Us* participant feedback, this response set was altered to add additional choices: 0 = I do not believe in God (or a higher power) or 0 = I am not religious. The scale is scored as the average of the items and is flagged as missing if more than two items do not have a valid response. Cronbach’s Alpha for the original scale is 0.91.^40^ Validity data were not presented in the original paper, though the Fetzer Institute^40^ report provides an extensive literature review.

**Religious Service Attendance.** One item on religious service attendance was obtained from the Women’s Health study.^32^ The wording of the item is “How often do you go to religious meetings or services?” The response options are: 5 = More than once a week; 4 = Once a week; 3 = 1 to 3 times per month; 2 = Less than once per month; 1 = Never (or almost never). In response to *All of Us* Participant feedback, a response option was added 0=I am not religious. There is no scoring for this item. Li et. al.^32^ showed that religious attendance was a protective factor for all cause mortality. Due to a transcription error, the initial response set for this item was incorrectly displayed for 11,795 participants in the version 7 data. These observations are flagged as ‘invalid’ in version 7 data and combined with the ‘PMI/Skip’ category. Users should use the ‘invalid’ flag to identify these observations and can apply missing data techniques in the analysis of these cases.
