## Appendix 4 for "Measuring Social Determinants of Health in the *All of Us* Research Program: Technical Document"

### Appendix 4: Item non-response and Cronbach’s Alpha for SDOH scales by participant characteristics

| **Appendix 4A: Item non-response and Cronbach's alphas by participant characteristic for loneliness, social support, instrumental social support, and emotional support scales (N=117,783)** | | | | | | | | |
| --- | --- | --- | --- | --- | --- | --- | --- | --- |
|  | **Loneliness** | | **Social support** | | **Instrumental social support** | | **Emotional social support** | |
|  | **Item  non-response** | **Alpha** | **Item  non-response** | **Alpha** | **Item  non-response** | **Alpha** | **Item  non-response** | **Alpha** |
| Total | 3018 (2.6%) | 0.87 | 3421 (2.9%) | 0.95 | 2088 (1.8%) | 0.95 | 2680 (2.3%) | 0.91 |
| RBR Overall | 433 (1.7%) | 0.87 | 285 (1.1%) | 0.94 | 150 (0.6%) | 0.96 | 227 (0.9%) | 0.90 |
| UBR Overall | 2768 (3.0%) | 0.87 | 3136 (3.4%) | 0.95 | 1938 (2.1%) | 0.95 | 2453 (2.7%) | 0.91 |
| Racial identity* |  |  |  |  |  |  |  |  |
| White | 1790 (2.0%) | 0.87 | 2105 (2.4%) | 0.95 | 1174 (1.3%) | 0.95 | 1597 (1.8%) | 0.91 |
| Black, African or African   American | 455 (5.1%) | 0.83 | 524 (5.8%) | 0.95 | 356 (4.0%) | 0.95 | 435 (4.8%) | 0.91 |
| Hispanic/Latino/Spanish | 457 (5.7%) | 0.85 | 435 (5.5%) | 0.95 | 319 (4.0%) | 0.95 | 362 (4.5%) | 0.91 |
| Asian | 58 (1.9%) | 0.87 | 69 (2.2%) | 0.95 | 45 (1.4%) | 0.95 | 58 (1.9%) | 0.92 |
| Native Hawaiian or   Pacific Islander (NHPI) | - | - | - | - | - | - | - | - |
| Middle Eastern or North   African (MENA) | - | - | - | - | - | - | - | - |
| Multi-Racial | 67 (1.6%) | 0.87 | 67 (1.6%) | 0.95 | 40 (1.0%) | 0.96 | 58 (1.4%) | 0.91 |
| None of these describe   me | 29 (2.8%) | 0.87 | 39 (3.8%) | 0.94 | 26 (2.5%) | 0.95 | 37 (3.6%) | 0.91 |
| Prefer not to answer or   skip | 148 (3.7%) | 0.87 | 159 (3.9%) | 0.95 | 108 (2.7%) | 0.95 | 120 (3.0%) | 0.91 |
| Sex assigned at birth |  |  |  |  |  |  |  |  |
| Female | 1839 (2.5%) | 0.87 | 2064 (2.8%) | 0.95 | 1189 (1.6%) | 0.95 | 1651 (2.2%) | 0.91 |
| Male | 1094 (2.7%) | 0.87 | 1256 (3.1%) | 0.95 | 833 (2.1%) | 0.96 | 937 (2.3%) | 0.91 |
| Intersex, none of these   describe me, prefer not   to answer, or skip | 85 (2.9%) | 0.87 | 100 (3.4%) | 0.95 | 66 (2.2%) | 0.95 | 91 (3.1%) | 0.91 |
| Gender identity |  |  |  |  |  |  |  |  |
| Woman | 1831 (2.5%) | 0.87 | 2069 (2.8%) | 0.95 | 1195 (1.6%) | 0.95 | 1655 (2.3%) | 0.91 |
| Man | 1092 (2.7%) | 0.87 | 1258 (3.1%) | 0.95 | 831 (2.1%) | 0.96 | 948 (2.4%) | 0.91 |
| Non-Binary | - | - | - | - | - | - | - | - |
| Transgender | - | - | - | - | - | - | - | - |
| None of these describe   me, prefer not to answer,   or skip | 91 (2.5%) | 0.88 | 87 (2.4%) | 0.95 | 58 (1.6%) | 0.95 | 73 (2.0%) | 0.91 |
| Sexual orientation |  |  |  |  |  |  |  |  |
| Straight | 2654 (2.6%) | 0.87 | 3032 (3.0%) | 0.95 | 1832 (1.8%) | 0.95 | 2370 (2.3%) | 0.91 |
| Lesbian | - | - | - | - | - | - | - | - |
| Gay | 55 (1.8%) | 0.88 | 61 (2.0%) | 0.95 | 40 (1.3%) | 0.96 | 47 (1.6%) | 0.91 |
| Bisexual | 50 (1.1%) | 0.86 | 69 (1.5%) | 0.94 | 42 (0.9%) | 0.95 | 51 (1.1%) | 0.91 |
| Multiple | - | - | - | - | - | - | - | - |
| None of these describe   me | 76 (3.2%) | 0.86 | 75 (3.2%) | 0.94 | 55 (2.4%) | 0.95 | 61 (2.7%) | 0.90 |
| Prefer not to answer or   skip | 150 (4.0%) | 0.87 | 152 (4.1%) | 0.95 | 102 (2.7%) | 0.95 | 124 (3.3%) | 0.91 |
| Educational attainment |  |  |  |  |  |  |  |  |
| College graduate or   advanced degree | 1374 (1.9%) | 0.87 | 1594 (2.2%) | 0.95 | 909  (1.2%) | 0.95 | 1212 (1.7%) | 0.91 |
| College (Years One to   Three) | 752 (2.7%) | 0.88 | 887 (3.2%) | 0.95 | 530 (1.9%) | 0.95 | 692 (2.5%) | 0.91 |
| Grade Twelve or GED | 517 (4.8%) | 0.86 | 564 (5.3%) | 0.95 | 376 (3.5%) | 0.95 | 470 (4.4%) | 0.91 |
| Less than high school   degree or equivalent | 221 (8.6%) | 0.83 | 218 (8.4%) | 0.94 | 154 (6.0%) | 0.93 | 185 (7.2%) | 0.91 |
| Prefer not to answer or   skip | 154 (4.5%) | 0.86 | 158 (4.6%) | 0.95 | 119 (3.5%) | 0.95 | 121 (3.5%) | 0.91 |
| Income |  |  |  |  |  |  |  |  |
| >150k | 261 (1.2%) | 0.86 | 352 (1.7%) | 0.94 | 173 (0.8%) | 0.95 | 265 (1.3%) | 0.90 |
| 100k-150k | 294 (1.6%) | 0.86 | 356 (1.9%) | 0.94 | 194 (1.0%) | 0.95 | 257 (1.4%) | 0.90 |
| 50k-100k | 682 (2.2%) | 0.86 | 774 (2.5%) | 0.94 | 453 (1.5%) | 0.95 | 568 (1.8%) | 0.90 |
| <50k | 1086 (3.3%) | 0.87 | 1227 (3.8%) | 0.95 | 762 (2.3%) | 0.95 | 1001 (3.1%) | 0.91 |
| Prefer not to answer or   skip | 695 (4.9%) | 0.86 | 712 (5.0%) | 0.95 | 506 (3.6%) | 0.95 | 589 (4.1%) | 0.91 |
| Disability |  |  |  |  |  |  |  |  |
| No disabilities identified | 1144 (2.2%) | 0.86 | 1292 (2.5%) | 0.95 | 793 (1.5%) | 0.96 | 995 (1.9%) | 0.91 |
| Living with disabilities | 500 (3.2%) | 0.87 | 569 (3.6%) | 0.94 | 355 (2.3%) | 0.95 | 442 (2.8%) | 0.91 |
| **NA | 1374 (2.7%) | 0.87 | 1560 (3.1%) | 0.95 | 939 (1.9%) | 0.95 | 1243 (2.5%) | 0.91 |
| Survey language |  |  |  |  |  |  |  | |
| English | 2743 (2.4%) | 0.87 | 3175 (2.8%) | 0.95 | 1908 (1.7%) | 0.95 | 2468 (2.1%) | 0.91 |
| Spanish | 275 (10.6%) | 0.81 | 245 (9.5%) | 0.94 | 179 (7.0%) | 0.93 | 212 (8.2%) | 0.90 |
| Notes: RBR: Represented in biomedical research; UBR: Underrepresented in biomedical research  -: Cells with counts below 20 were suppressed; counts were also suppressed to prevent participant re-identification due to small sample sizes (NHPI, MENA, Non-Binary, Transgender, Lesbian, and Multiple Sexual Orientation) *Racial and Ethnic Identity: Racial Identity (Asian, H/L/S, White, etc.); Ethnic Identity (Japanese, Cuban, Irish, etc.). Ethnic Identity data not provided due to space limitations.  **Participants that responded to the Basics survey before October 22, 2019 do not have demographic information on disability | | | | | | | | |

| **Appendix 4B: Item non-response and Cronbach's alphas by participant characteristic for perceived stress, everyday discrimination, discrimination in healthcare settings, and social cohesion scales (N=117,783)** | | | | | | | | | |
| --- | --- | --- | --- | --- | --- | --- | --- | --- | --- |
|  | **Perceived stress** | | | **Everyday discrimination** | | **Discrimination in healthcare settings** | | **Social cohesion** | |
|  | **Item  non-response** | **Alpha** | **Item  non-response** | | **Alpha** | **Item  non-response** | **Alpha** | **Item  non-response** | **Alpha** |
| Total | 7129 (6.1%) | 0.91 | 3915 (3.3%) | | 0.91 | 2885 (2.5%) | 0.86 | 4770 (4.1%) | 0.87 |
| RBR Overall | 693 (2.7%) | 0.90 | 314 (1.2%) | | 0.89 | 267 (1.1%) | 0.89 | 145 (0.6%) | 0.86 |
| UBR Overall | 6436 (7.0%) | 0.91 | 3601 (3.9%) | | 0.91 | 2618 (2.8%) | 0.90 | 1731 (1.9%) | 0.87 |
| Racial identity* |  |  |  | |  |  |  |  |  |
| White | 4887 (5.6%) | 0.91 | 2427 (2.8%) | | 0.90 | 1685 (1.9%) | 0.89 | 3270 (3.7%) | 0.87 |
| Black, African or African   American | 843 (9.4%) | 0.87 | 577 (6.4%) | | 0.92 | 441 (4.9%) | 0.91 | 551 (6.1%) | 0.86 |
| Hispanic/Latino/Spanish | 660 (8.3%) | 0.87 | 499 (6.3%) | | 0.91 | 433 (5.4%) | 0.90 | 484 (6.1%) | 0.87 |
| Asian | 143 (4.6%) | 0.89 | 73 (2.3%) | | 0.91 | 53 (1.7%) | 0.90 | 75 (2.4%) | 0.86 |
| Native Hawaiian or   Pacific Islander (NHPI) | - | - | - | | - | - | - | - | - |
| Middle Eastern or North   African (MENA) | - | - | - | | - | - | - | - | - |
| Multi-Racial | 156 (3.7%) | 0.91 | 89 (2.1%) | | 0.92 | 73 (1.7%) | 0.90 | 99 (2.4%) | 0.86 |
| None of these describe   me | 72 (6.9%) | 0.91 | 53 (5.1%) | | 0.92 | 38 (3.7%) | 0.90 | 55 (5.3%) | 0.88 |
| Prefer not to answer or   skip | 336 (8.3%) | 0.91 | 184 (4.6%) | | 0.91 | 139 (3.5%) | 0.90 | 215 (5.3%) | 0.86 |
| Sex assigned at birth |  |  |  | |  |  |  |  |  |
| Female | 4359 (5.9%) | 0.91 | 2463 (3.3%) | | 0.91 | 1814 (2.4%) | 0.89 | 2860 (3.8%) | 0.87 |
| Male | 2571 (6.4%) | 0.89 | 1324 (3.3%) | | 0.92 | 970 (2.4%) | 0.90 | 1758 (4.4%) | 0.86 |
| Intersex, none of these   describe me, prefer not   to answer, or skip | 199 (6.7%) | 0.91 | 128 (4.3%) | | 0.92 | 100 (3.4%) | 0.89 | 151 (5.1%) | 0.87 |
| Gender identity |  |  |  | |  |  |  |  |  |
| Woman | 4337 (5.9%) | 0.91 | 2464 (3.4%) | | 0.91 | 1803 (2.5%) | 0.89 | 2842 (3.9%) | 0.87 |
| Man | 2563 (6.4%) | 0.89 | 1327 (3.3%) | | 0.91 | 977 (2.4%) | 0.90 | 1757 (4.4%) | 0.86 |
| Non-Binary | - | - | - | | - | - | - | - | - |
| Transgender | - | - | - | | - | - | - | - | - |
| None of these describe   me, prefer not to answer,   or skip | 214 (6.0%) | 0.92 | 113 (3.2%) | | 0.93 | 97 (2.7%) | 0.91 | 153 (4.3%) | 0.86 |
| Sexual orientation |  |  |  | |  |  |  |  |  |
| Straight | 6385 (6.3%) | 0.90 | 3461 (3.4%) | | 0.91 | 2536 (2.5%) | 0.89 | 4244 (4.2%) | 0.87 |
| Lesbian | - | - | - | | - | - | - | - | - |
| Gay | 137 (4.5%) | 0.91 | 62 (2.0%) | | 0.92 | 41 (1.3%) | 0.90 | 107 (3.5%) | 0.85 |
| Bisexual | 140 (3.0%) | 0.92 | 78 (1.7%) | | 0.90 | 66 (1.4%) | 0.90 | 89 (1.9%) | 0.85 |
| Multiple | - | - | - | | - | - | - | - | - |
| None of these describe   me | 98 (4.2%) | 0.91 | 88 (3.8%) | | 0.92 | 66 (2.8%) | 0.91 | 77 (3.3%) | 0.83 |
| Prefer not to answer or   skip | 288 (7.7%) | 0.90 | 186 (5.0%) | | 0.92 | 149 (4.0%) | 0.90 | 195 (5.2%) | 0.86 |
| Educational attainment |  |  |  | |  |  |  |  |  |
| College graduate or   advanced degree | 3865 (5.3%) | 0.91 | 1790 (2.4%) | | 0.90 | 1261 (1.7%) | 0.89 | 2519 (3.4%) | 0.86 |
| College (Years One to   Three) | 1700 (6.2%) | 0.91 | 1021 (3.7%) | | 0.91 | 754 (2.8%) | 0.90 | 1189 (4.3%) | 0.88 |
| Grade Twelve or GED | 1000 (9.4%) | 0.89 | 683 (6.4%) | | 0.92 | 508 (4.8%) | 0.90 | 651 (6.1%) | 0.88 |
| Less than high school   degree or equivalent | 301 (11.6%) | 0.84 | 249 (9.7%) | | 0.92 | 212 (8.2%) | 0.90 | 234 (9.1%) | 0.87 |
| Prefer not to answer or   skip | 263 (7.6%) | 0.90 | 171 (5.0%) | | 0.91 | 149 (4.3%) | 0.89 | 177 (5.1%) | 0.87 |
| Income |  |  |  | |  |  |  |  |  |
| >150k | 894 (4.2%) | 0.89 | 336 (1.6%) | | 0.89 | 256 (1.2%) | 0.89 | 575 (2.7%) | 0.85 |
| 100k-150k | 835 (4.5%) | 0.90 | 368 (2.0%) | | 0.89 | 260 (1.4%) | 0.89 | 601 (3.2%) | 0.85 |
| 50k-100k | 1768 (5.7%) | 0.91 | 872 (2.8%) | | 0.89 | 614 (2.0%) | 0.90 | 1150 (3.7%) | 0.86 |
| <50k | 2320 (7.1%) | 0.90 | 1456 (4.5%) | | 0.90 | 1058 (3.2%) | 0.89 | 1580 (4.8%) | 0.87 |
| Prefer not to answer or   skip | 1312 (9.2%) | 0.90 | 883 (6.2%) | | 0.90 | 697 (4.9%) | 0.90 | 864 (6.1%) | 0.87 |
| Disability |  |  |  | |  |  |  |  |  |
| No disabilities identified | 2821 (5.4%) | 0.90 | 1424 (2.7%) | | 0.90 | 1087 (2.1%) | 0.89 | 1820 (3.5%) | 0.87 |
| Living with disabilities | 1102 (7.0%) | 0.91 | 670 (4.3%) | | 0.91 | 483 (3.1%) | 0.90 | 793 (5.1%) | 0.87 |
| **NA | 3206 (6.4%) | 0.91 | 1821 (3.6%) | | 0.91 | 1315 (2.6%) | 0.90 | 2157 (4.3%) | 0.86 |
| Survey language |  |  |  | |  |  |  |  | |
| English | 6805 (5.9%) | 0.91 | 3617 (3.1%) | | 0.87 | 2651 (2.3%) | 0.90 | 4495 (3.9%) | 0.87 |
| Spanish | 324 (12.5%) | 0.86 | 298 (11.5%) | | 0.83 | 234 (9.1%) | 0.88 | 275 (10.6%) | 0.86 |
| Notes: RBR: Represented in biomedical research; UBR: Underrepresented in biomedical research  -: Cells with counts below 20 were suppressed; counts were also suppressed to prevent participant re-identification due to small sample sizes (NHPI, MENA, Non-Binary, Transgender, Lesbian, and Multiple Sexual Orientation) *Racial and Ethnic Identity: Racial Identity (Asian, H/L/S, White, etc.); Ethnic Identity (Japanese, Cuban, Irish, etc.). Ethnic Identity data not provided due to space limitations.  **Participants that responded to the Basics survey before October 22, 2019 do not have demographic information on disability | | | | | | | | | |

| **Appendix 4C: Item non-response and Cronbach's alphas by participant characteristic for neighborhood physical disorder, neighborhood physical disorder, daily spiritual experiences, and PANES - Walking and bicycling scales (N=117,783)** | | | | | | | | | |
| --- | --- | --- | --- | --- | --- | --- | --- | --- | --- |
|  | **Neighborhood physical disorder** | | **Neighborhood social disorder** | | | **Daily spiritual experiences** | | **PANES - Walking and Bicycling** | |
|  | **Item  non-response** | **Alpha** | **Item  non-response** | **Alpha** | **Item  non-response** | | **Alpha** | **Item  non-response** | **Alpha** |
| Total | 3417 (2.9%) | 0.84 | 6215 (5.3%) | 0.87 | 1334 (1.1%) | | 0.81 | 5771 (4.9%) | 0.78 |
| RBR Overall | 263 (1.0%) | 0.83 | 567 (2.2%) | 0.85 | 143 (0.6%) | | 0.81 | 657 (2.6%) | 0.80 |
| UBR Overall | 3153 (3.4%) | 0.84 | 5648 (6.1%) | 0.87 | 1191 (1.3%) | | 0.82 | 5114 (5.5%) | 0.78 |
| Racial identity* |  |  |  |  |  | |  |  |  |
| White | 2157 (2.5%) | 0.82 | 4139 (4.7%) | 0.86 | 684 (0.8%) | | 0.81 | 3898 (4.4%) | 0.79 |
| Black, African or African   American | 446 (5.0%) | 0.84 | 763 (8.5%) | 0.88 | 248 (2.8%) | | 0.87 | 702 (7.8%) | 0.74 |
| Hispanic/Latino/Spanish | 492 (6.2%) | 0.84 | 676 (8.5%) | 0.87 | 217 (2.7%) | | 0.84 | 552 (6.9%) | 0.73 |
| Asian | 58 (1.9%) | 0.85 | 105 (3.4%) | 0.86 | 39 (1.3%) | | 0.79 | 118 (3.8%) | 0.70 |
| Native Hawaiian or   Pacific Islander (NHPI) | - | - | - | - | - | | - | - | - |
| Middle Eastern or North   African (MENA) | - | - | - | - | - | | - | - | - |
| Multi-Racial | 66 (1.6%) | 0.85 | 124 (2.9%) | 0.87 | 47 (1.1%) | | 0.79 | 159 (3.8%) | 0.74 |
| None of these describe   me | 30 (2.9%) | 0.84 | 72 (6.9%) | 0.88 | 22 (2.1%) | | 0.81 | 55 (5.3%) | 0.78 |
| Prefer not to answer or   skip | 152 (3.8%) | 0.84 | 304 (7.5%) | 0.87 | 63 (1.6%) | | 0.81 | 256 (6.4%) | 0.78 |
| Sex assigned at birth |  |  |  |  |  | |  |  |  |
| Female | 2156 (2.9%) | 0.84 | 3936 (5.3%) | 0.87 | 784 (1.1%) | | 0.81 | 3682 (5.0%) | 0.79 |
| Male | 1158 (2.9%) | 0.83 | 2088 (5.2%) | 0.87 | 506 (1.3%) | | 0.82 | 1903 (4.7%) | 0.78 |
| Intersex, none of these   describe me, prefer not   to answer, or skip | 103 (3.5%) | 0.85 | 191 (6.4%) | 0.87 | 44 (1.5%) | | 0.81 | 186 (6.3%) | 0.78 |
| Gender identity |  |  |  |  |  | |  |  |  |
| Woman | 2143 (2.9%) | 0.84 | 3898 (5.3%) | 0.87 | 775 (1.1%) | | 0.81 | 3652 (5.0%) | 0.79 |
| Man | 1160 (2.9%) | 0.83 | 2084 (5.2%) | 0.87 | 509 (1.3%) | | 0.82 | 1893 (4.7%) | 0.78 |
| Non-Binary | - | - | - | - | - | | - | - | - |
| Transgender | - | - | - | - | - | | - | - | - |
| None of these describe   me, prefer not to answer,   or skip | 109 (3.1%) | 0.85 | 218 (6.1%) | 0.87 | 46 (1.3%) | | 0.79 | 195 (5.5%) | 0.77 |
| Sexual orientation |  |  |  |  |  | |  |  |  |
| Straight | 3017 (3.0%) | 0.83 | 5506 (5.4%) | 0.87 | 1159 (1.1%) | | 0.82 | 4940 (4.9%) | 0.79 |
| Lesbian | - | - | - | - | - | | - | - | - |
| Gay | 66 (2.2%) | 0.84 | 125 (4.1%) | 0.87 | 20 (0.7%) | | 0.79 | 146 (4.8%) | 0.77 |
| Bisexual | 65 (1.4%) | 0.86 | 112 (2.4%) | 0.87 | 32 (0.7%) | | 0.77 | 181 (3.9%) | 0.78 |
| Multiple | - | - | - | - | - | | - | - | - |
| None of these describe   me | 76 (3.2%) | 0.83 | 116 (5.0%) | 0.85 | 30 (1.3%) | | 0.77 | 138 (5.9%) | 0.77 |
| Prefer not to answer or   skip | 156 (4.2%) | 0.84 | 281 (7.5%) | 0.87 | 80 (2.1%) | | 0.81 | 275 (7.3%) | 0.77 |
| Educational attainment |  |  |  |  |  | |  |  |  |
| College graduate or   advanced degree | 1541 (2.1%) | 0.83 | 3207 (4.4%) | 0.85 | 535 (0.7%) | | 0.80 | 2917 (4.0%) | 0.78 |
| College (Years One to   Three) | 915 (3.3%) | 0.84 | 1615 (5.9%) | 0.88 | 330 (1.2%) | | 0.83 | 1490 (5.4%) | 0.79 |
| Grade Twelve or GED | 563 (5.3%) | 0.84 | 847 (7.9%) | 0.88 | 275 (2.6%) | | 0.85 | 837 (7.8%) | 0.80 |
| Less than high school   degree or equivalent | 245 (9.5%) | 0.81 | 299 (11.6%) | 0.86 | 109 (4.2%) | | 0.86 | 280 (10.8%) | 0.80 |
| Prefer not to answer or   skip | 152 (4.4%) | 0.85 | 246 (7.2%) | 0.87 | 85 (2.5%) | | 0.82 | 246 (7.2%) | 0.78 |
| Income |  |  |  |  |  | |  |  |  |
| >150k | 303 (1.4%) | 0.82 | 695 (3.3%) | 0.83 | 94 (0.5%) | | 0.80 | 585 (2.8%) | 0.79 |
| 100k-150k | 328 (1.8%) | 0.81 | 714 (3.8%) | 0.84 | 89 (0.5%) | | 0.81 | 667 (3.6%) | 0.78 |
| 50k-100k | 768 (2.5%) | 0.82 | 1464 (4.7%) | 0.85 | 229 (0.7%) | | 0.82 | 1315 (4.3%) | 0.79 |
| <50k | 1267 (3.9%) | 0.84 | 2131 (6.5%) | 0.88 | 474 (1.5%) | | 0.82 | 2039 (6.2%) | 0.78 |
| Prefer not to answer or   skip | 751 (5.3%) | 0.83 | 1211 (8.5%) | 0.87 | 447 (3.1%) | | 0.83 | 1165 (8.2%) | 0.78 |
| Disability |  |  |  |  |  | |  |  |  |
| No disabilities identified | 1246 (2.4%) | 0.84 | 2283 (4.4%) | 0.87 | 577 (1.1%) | | 0.81 | 2210 (4.3%) | 0.79 |
| Living with disabilities | 596 (3.8%) | 0.83 | 977 (6.2%) | 0.88 | 197 (1.3%) | | 0.82 | 942 (6.0%) | 0.78 |
| **NA | 1574 (3.1%) | 0.83 | 2955 (5.9%) | 0.87 | 560 (1.1%) | | 0.81 | 2619 (5.2%) | 0.78 |
| Survey language |  |  |  |  |  | |  |  | |
| English | 3108 (2.7%) | 0.83 | 5813 (5.1%) | 0.87 | 1241 (1.1%) | | 0.81 | 5495 (4.8%) | 0.79 |
| Spanish | 309 (11.9%) | 0.80 | 402 (15.5%) | 0.83 | 93 (3.6%) | | 0.85 | 276 (10.7%) | 0.74 |
| Notes: PANES: Physical Activity and Neighborhood Environment Scale. For PANES items related to walking and bicycling, respondents who answered this item “does not apply to my neighborhood” were not included in score distributions and were not counted in non-response totals.  RBR: Represented in biomedical research; UBR: Underrepresented in biomedical research  -: Cells with counts below 20 were suppressed; counts were also suppressed to prevent participant re-identification due to small sample sizes (NHPI, MENA, Non-Binary, Transgender, Lesbian, and Multiple Sexual Orientation) *Racial and Ethnic Identity: Racial Identity (Asian, H/L/S, White, etc.); Ethnic Identity (Japanese, Cuban, Irish, etc.). Ethnic Identity data not provided due to space limitations.  **Participants that responded to the Basics survey before October 22, 2019 do not have demographic information on disability | | | | | | | | | |

| **Appendix 4D: Item non-response by participant characteristic for PANES - Crime and safety, food insecurity, housing instability, housing quality, and religious service attendance measures (N=117,783)** | | | | | |
| --- | --- | --- | --- | --- | --- |
|  | **PANES - Crime and Safety** | **Food insecurity** | **Housing instability** | **Housing quality** | **Religious service attendance*** |
| Total | 15,293 (13.0%) | 1542 (1.3%) | 3620 (3.1%) | 6134 (5.2%) | 1825 (1.5%) |
| RBR Overall | 2040 (8.0%) | 127 (0.5%) | 327 (1.3%) | 651 (2.6%) | 180 (0.7%) |
| UBR Overall | 13,253 (14.4%) | 1415 (1.5%) | 3293 (3.6%) | 5483 (5.9%) | 1645 (1.8%) |
| Racial identity* |  |  |  |  |  |
| White | 10,636 (12.1%) | 775 (0.9%) | 2052 (2.3%) | 4664 (5.3%) | 973 (1.1%) |
| Black, African or African   American | 1617 (18.0%) | 295 (3.3%) | 593 (6.6%) | 419 (4.7%) | 332 (3.7%) |
| Hispanic/Latino/Spanish | 1363 (17.1%) | 258 (3.2%) | 556 (7.0%) | 412 (5.2%) | 283 (3.6%) |
| Asian | 347 (11.1%) | 43 (1.4%) | 90 (2.9%) | 126 (4.0%) | 50 (1.6%) |
| Native Hawaiian or   Pacific Islander (NHPI) | - | - | - | - | - |
| Middle Eastern or North   African (MENA) | - | - | - | - | - |
| Multi-Racial | 443 (10.5%) | 52 (1.2%) | 103 (2.4%) | 141 (3.3%) | 58 (1.4%) |
| None of these describe   me | 171 (16.5%) | - | 48 (4.6%) | 50 (4.8%) | 31 (3.0%) |
| Prefer not to answer or   skip | 637 (15.8%) | 85 (2.1%) | 149 (3.7%) | 295 (7.3%) | 84 (2.1%) |
| Sex assigned at birth |  |  |  |  |  |
| Female | 9902 (13.3%) | 908 (1.2%) | 2085 (2.8%) | 3594 (4.8%) | 1101 (1.5%) |
| Male | 4998 (12.4%) | 575 (1.4%) | 1435 (3.6%) | 2367 (5.9%) | 658 (1.6%) |
| Intersex, none of these   describe me, prefer not   to answer, or skip | 393 (13.2%) | 59 (2.0%) | 100 (3.4%) | 173 (5.8%) | 66 (2.2%) |
| Gender identity |  |  |  |  |  |
| Woman | 9814 (13.4%) | 901 (1.2%) | 2079 (2.8%) | 3574 (4.9%) | 1094 (1.5%) |
| Man | 4981 (12.5%) | 580 (1.5%) | 1426 (3.6%) | 2365 (5.9%) | 662 (1.7%) |
| Non-Binary | - | - | - | - | - |
| Transgender | - | - | - | - | - |
| None of these describe   me, prefer not to answer,   or skip | 433 (12.1%) | 56 (1.6%) | 109 (3.0%) | 182 (5.1%) | 63 (1.8%) |
| Sexual orientation |  |  |  |  |  |
| Straight | 13493 (13.2%) | 1326 (1.3%) | 3183 (3.1%) | 5525 (5.4%) | 1578 (1.6%) |
| Lesbian | - | - | - | - | - |
| Gay | 321 (10.5%) | 28 (0.9%) | 60 (2.0%) | 113 (3.7%) | 32 (1.1%) |
| Bisexual | 403 (8.7%) | 38 (0.8%) | 72 (1.6%) | 136 (2.9%) | 47 (1.0%) |
| Multiple | - | - | - | - | - |
| None of these describe   me | 302 (12.9%) | 60 (1.9%) | 97 (3.1%) | 71 (3.0%) | 44 (1.9%) |
| Prefer not to answer or   skip | 552 (14.7%) | 86 (2.0%) | 203 (4.7%) | 224 (6.0%) | 104 (2.8%) |
| Educational attainment |  |  |  |  |  |
| College graduate or   advanced degree | 8671 (11.8%) | 604 (0.8%) | 1581 (2.2%) | 3835 (5.2%) | 743 (1.0%) |
| College (Years One to   Three) | 3774 (13.8%) | 388 (1.4%) | 932 (3.4%) | 1331 (4.9%) | 474 (1.7%) |
| Grade Twelve or GED | 1832 (17.1%) | 327 (3.1%) | 643 (5.9%) | 619 (5.8%) | 364 (3.4%) |
| Less than high school   degree or equivalent | 551 (21.3%) | 129 (5.0%) | 292 (11.3%) | 139 (5.4%) | 137 (5.3%) |
| Prefer not to answer or   skip | 465 (13.5%) | 94 (2.7%) | 181 (5.2%) | 210 (6.1%) | 107 (3.1%) |
| Income |  |  |  |  |  |
| >150k | 2108 (9.9%) | 106 (0.5%) | 338 (1.6%) | 995 (4.7%) | 137 (0.7%) |
| 100k-150k | 1942 (10.4%) | 110 (0.6%) | 305 (1.6%) | 844 (4.5%) | 151 (0.8%) |
| 50k-100k | 3897 (12.6%) | 273 (0.9%) | 752 (2.4%) | 1663 (5.4%) | 342 (1.1%) |
| <50k | 4895 (15.0%) | 606 (1.9%) | 1340 (4.1%) | 1616 (4.9%) | 687 (2.1%) |
| Prefer not to answer or   skip | 2451 (17.2%) | 447 (3.1%) | 885 (6.2%) | 1016 (7.1%) | 508 (3.6%) |
| Disability |  |  |  |  |  |
| No disabilities identified | 6277 (12.1%) | 641 (1.2%) | 1284 (2.5%) | 2439 (4.7%) | 728 (1.4%) |
| Living with disabilities | 2253 (14.4%) | 282 (1.8%) | 589 (3.8%) | 744 (4.8%) | 326 (2.1%) |
| **NA | 6763 (13.5%) | 619 (1.2%) | 1747 (3.5%) | 2951 (5.9%) | 771 (1.5%) |
| Survey language |  |  |  |  |  |
| English | 14630 (12.7%) | 1399 (1.2%) | 3296 (2.9%) | 5955 (5.2%) | 1703 (1.5%) |
| Spanish | 662 (25.6%) | 143 (5.5%) | 324 (12.5%) | 179 (6.9%) | 122 (4.7%) |
| Notes: PANES: Physical Activity and Neighborhood Environment Scale. For PANES items related to walking and bicycling, respondents who answered this item “does not apply to my neighborhood” were not included in score distributions and were not counted in non-response totals.  RBR: Represented in biomedical research; UBR: Underrepresented in biomedical research  -: Cells with counts below 20 were suppressed; counts were also suppressed to prevent participant re-identification due to small sample sizes (NHPI, MENA, Non-Binary, Transgender, Lesbian, and Multiple Sexual Orientation) *Participants who responded to the incorrect response set for the Religious Service Attendance item (N=11,795) are flagged as ‘invalid’ in version 7 data; these respondents are not included in item non-response calculations. **Racial and Ethnic Identity: Racial Identity (Asian, H/L/S, White, etc.); Ethnic Identity (Japanese, Cuban, Irish, etc.). Ethnic Identity data not provided due to space limitations.  ***Participants that responded to the Basics survey before October 22, 2019 do not have demographic information on disability | | | | | |
