## Appendix 5 for "Measuring Social Determinants of Health in the *All of Us* Research Program: Technical Document"

### Appendix 5: Forest plots of multivariable logistic regression models predicting missingness within SDOH scales

**Figure A1. Odds of item non-response or incalculable score for the Loneliness Scale.**

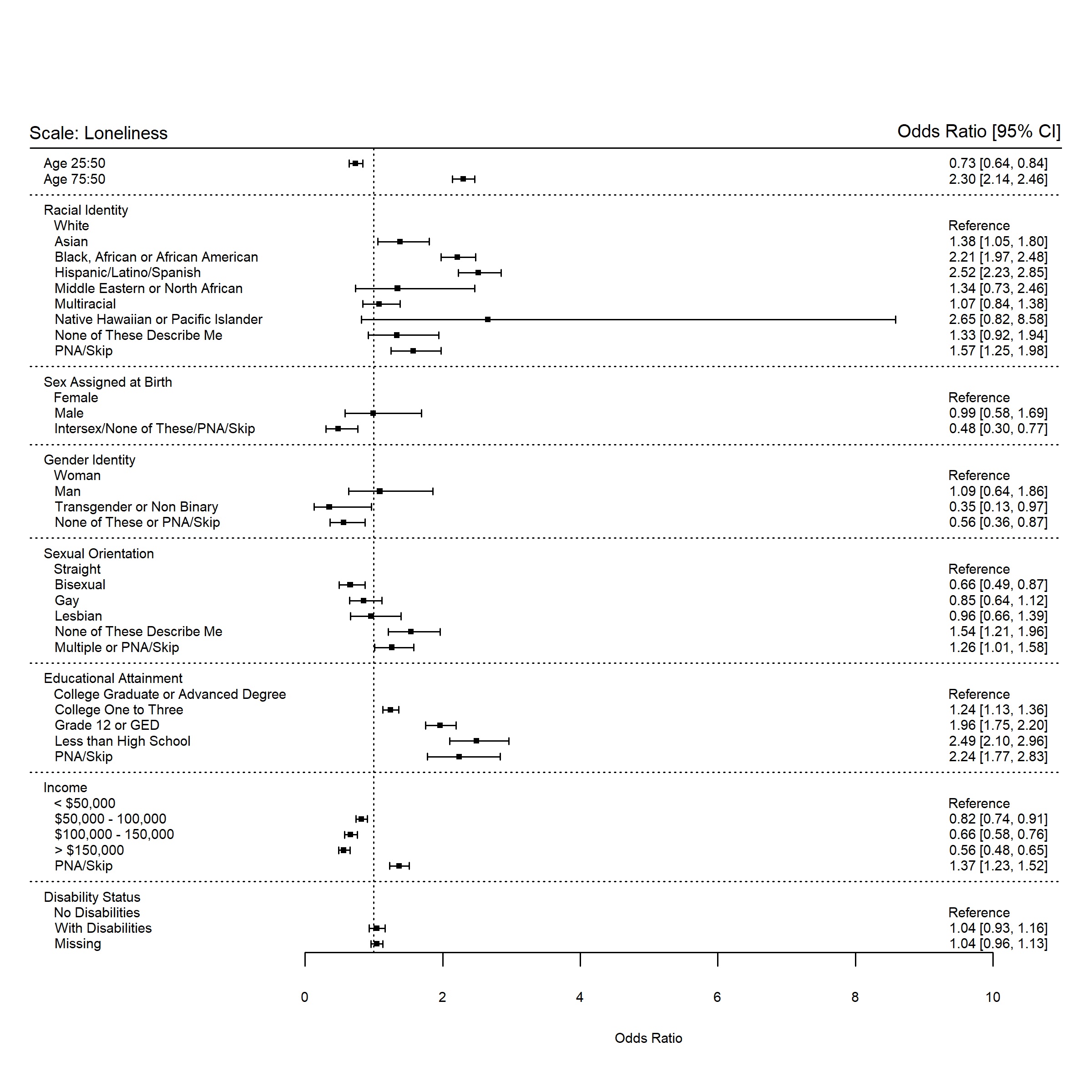

Notes: White; Black: Black, African or African-American; Hispanic: Hispanic/Latino/Spanish;

MENA: Middle Eastern or North African; NHPI: Native Hawaiian or Pacific Islander; PNA: Prefer not to answer

Sex at Birth: Sex Assigned at Birth

**Figure A2: Odds of item non-response or incalculable score for the Neighborhood Physical Disorder Scale.**

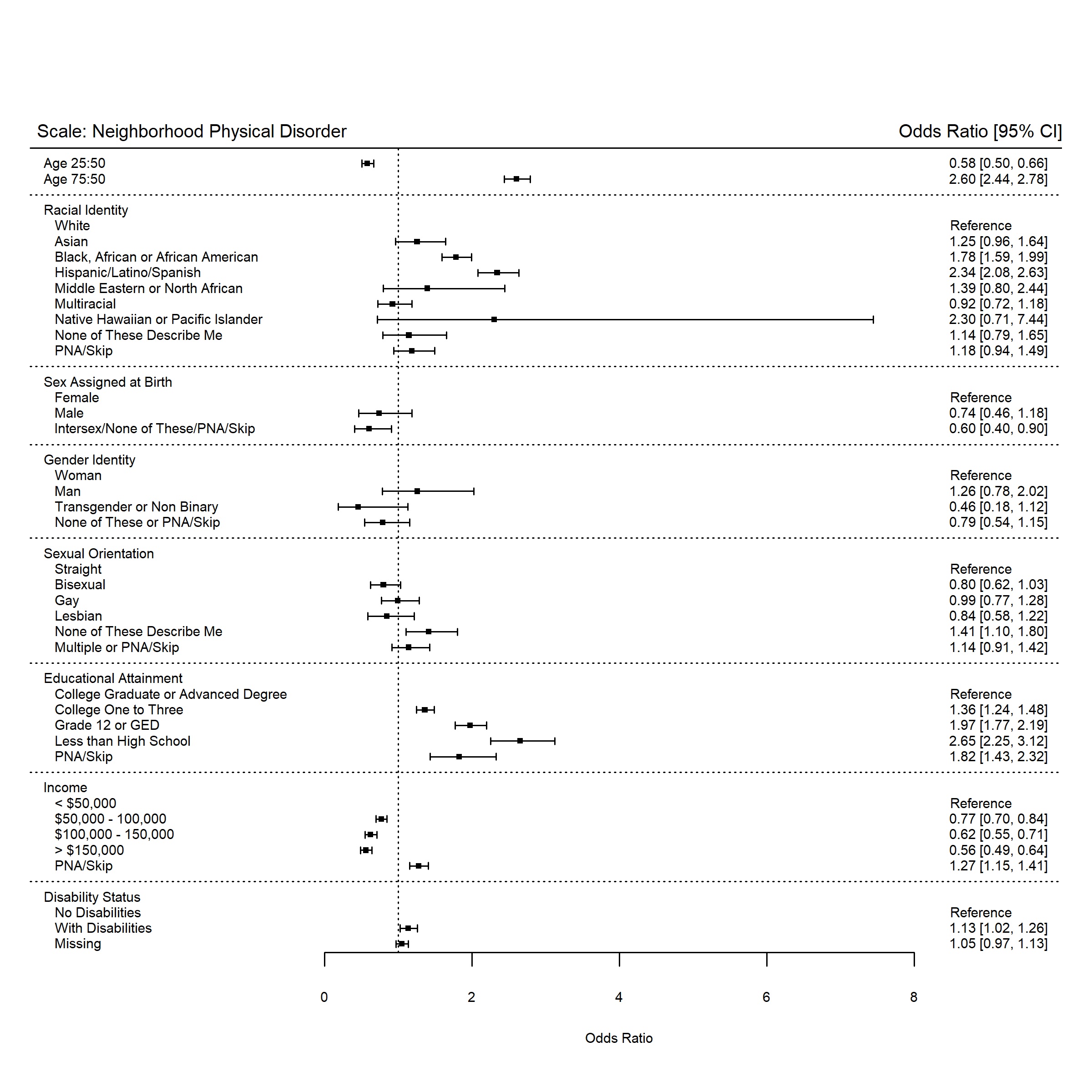

Notes: White; Black: Black, African or African-American; Hispanic: Hispanic/Latino/Spanish;

MENA: Middle Eastern or North African; NHPI: Native Hawaiian or Pacific Islander; PNA: Prefer not to answer

Sex at Birth: Sex Assigned at Birth

**Figure A3: Odds of item non-response or incalculable score for the Neighborhood Social Disorder Scale.**

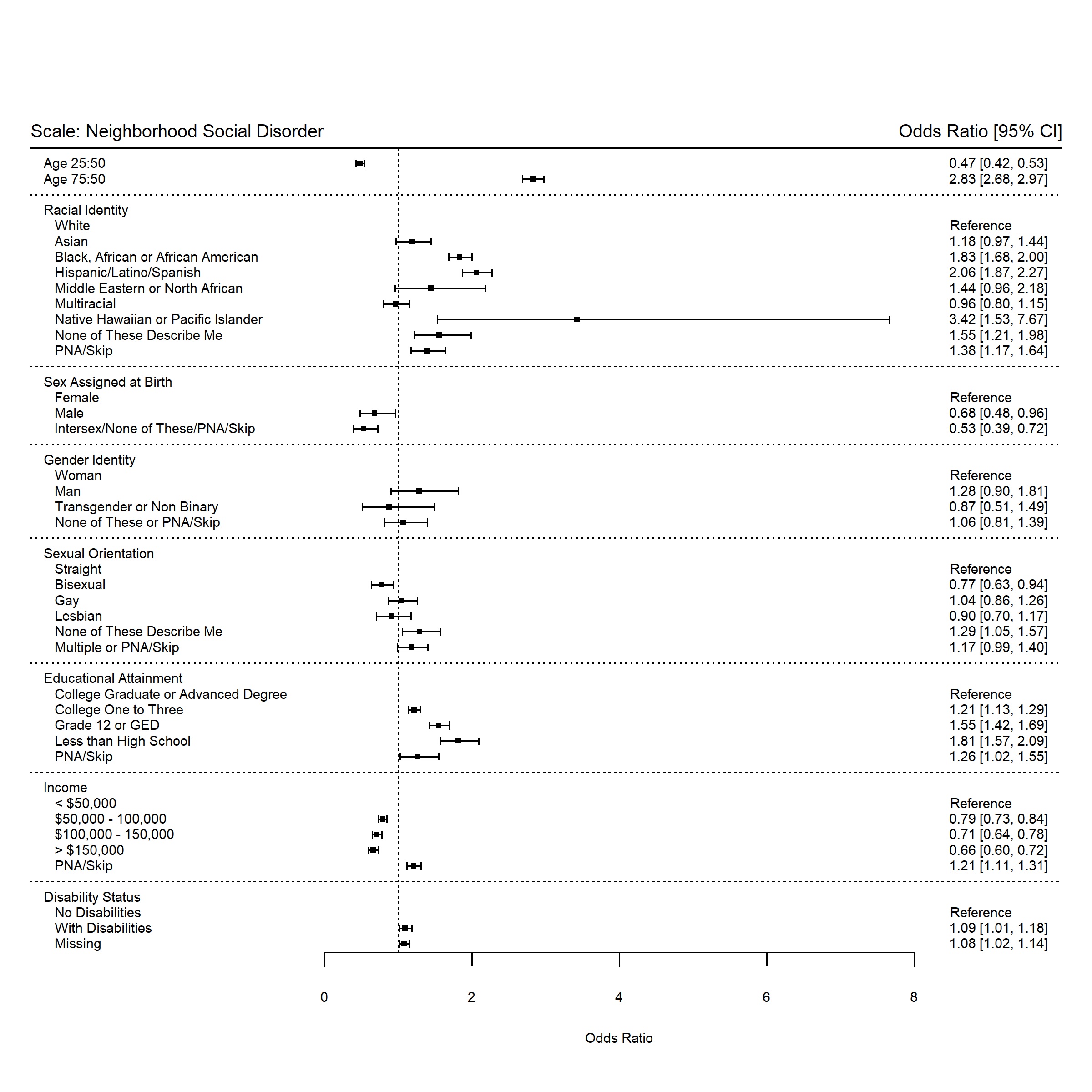

Notes: White; Black: Black, African or African-American; Hispanic: Hispanic/Latino/Spanish;

MENA: Middle Eastern or North African; NHPI: Native Hawaiian or Pacific Islander; PNA: Prefer not to answer

Sex at Birth: Sex Assigned at Birth

**Figure A4: Odds of item non-response or incalculable score for the Everyday Discrimination Scale.**

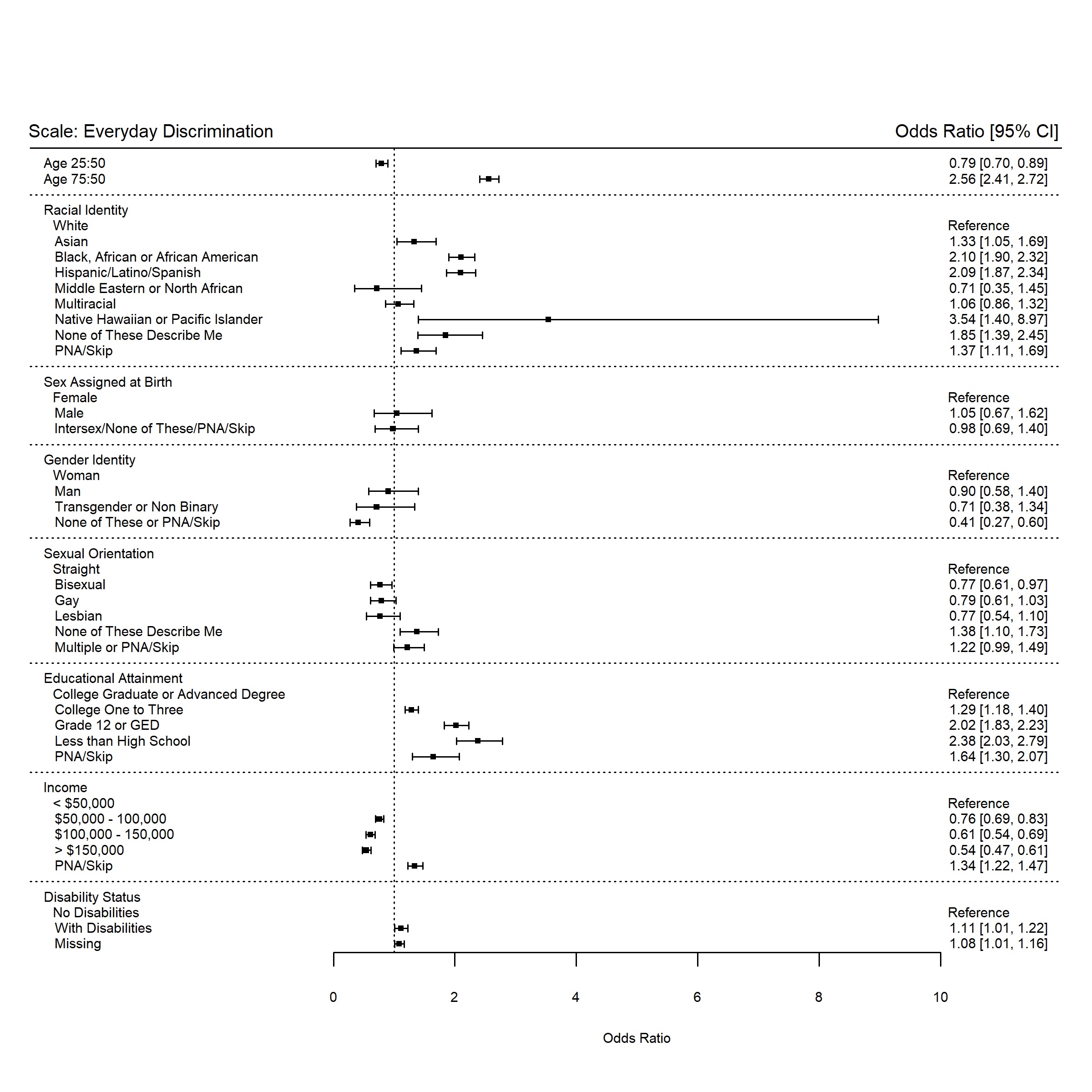

Notes: White; Black: Black, African or African-American; Hispanic: Hispanic/Latino/Spanish;

MENA: Middle Eastern or North African; NHPI: Native Hawaiian or Pacific Islander; PNA: Prefer not to answer

Sex at Birth: Sex Assigned at Birth

**Figure A5: Odds of item non-response or incalculable score for the Discrimination in Medical Settings Scale.**

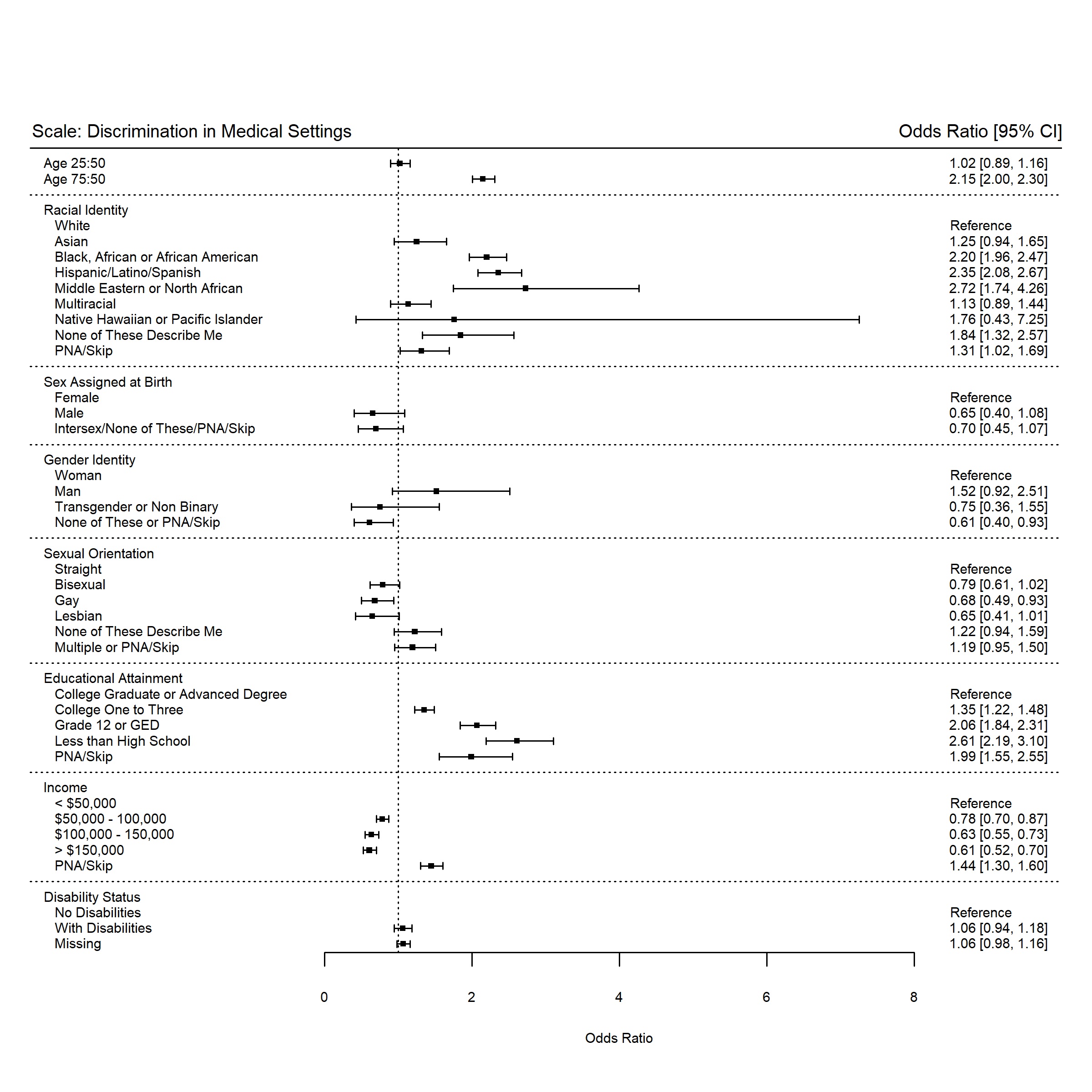

Notes: White; Black: Black, African or African-American; Hispanic: Hispanic/Latino/Spanish;

MENA: Middle Eastern or North African; NHPI: Native Hawaiian or Pacific Islander; PNA: Prefer not to answer

Sex at Birth: Sex Assigned at Birth

**Figure A6: Odds of item non-response or incalculable score for the Social Cohesion Scale.**

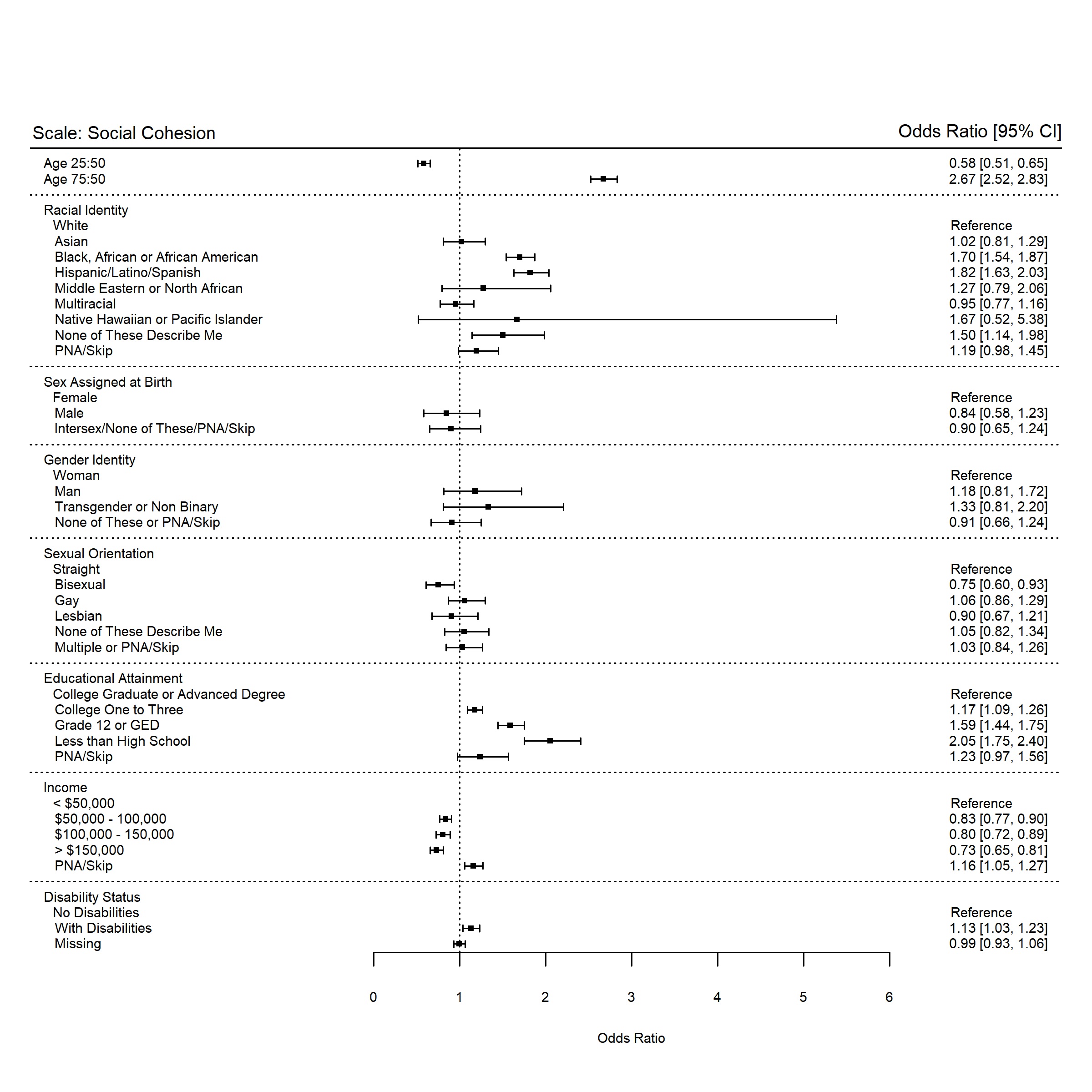

Notes: White: White; Black: Black, African or African-American; Hispanic: Hispanic/Latino/Spanish;

MENA: Middle Eastern or North African; NHPI: Native Hawaiian or Pacific Islander; PNA: Prefer not to answer

Sex at Birth: Sex Assigned at Birth

**Figure A7: Odds of item non-response or incalculable score for the Perceived Stress Scale.**

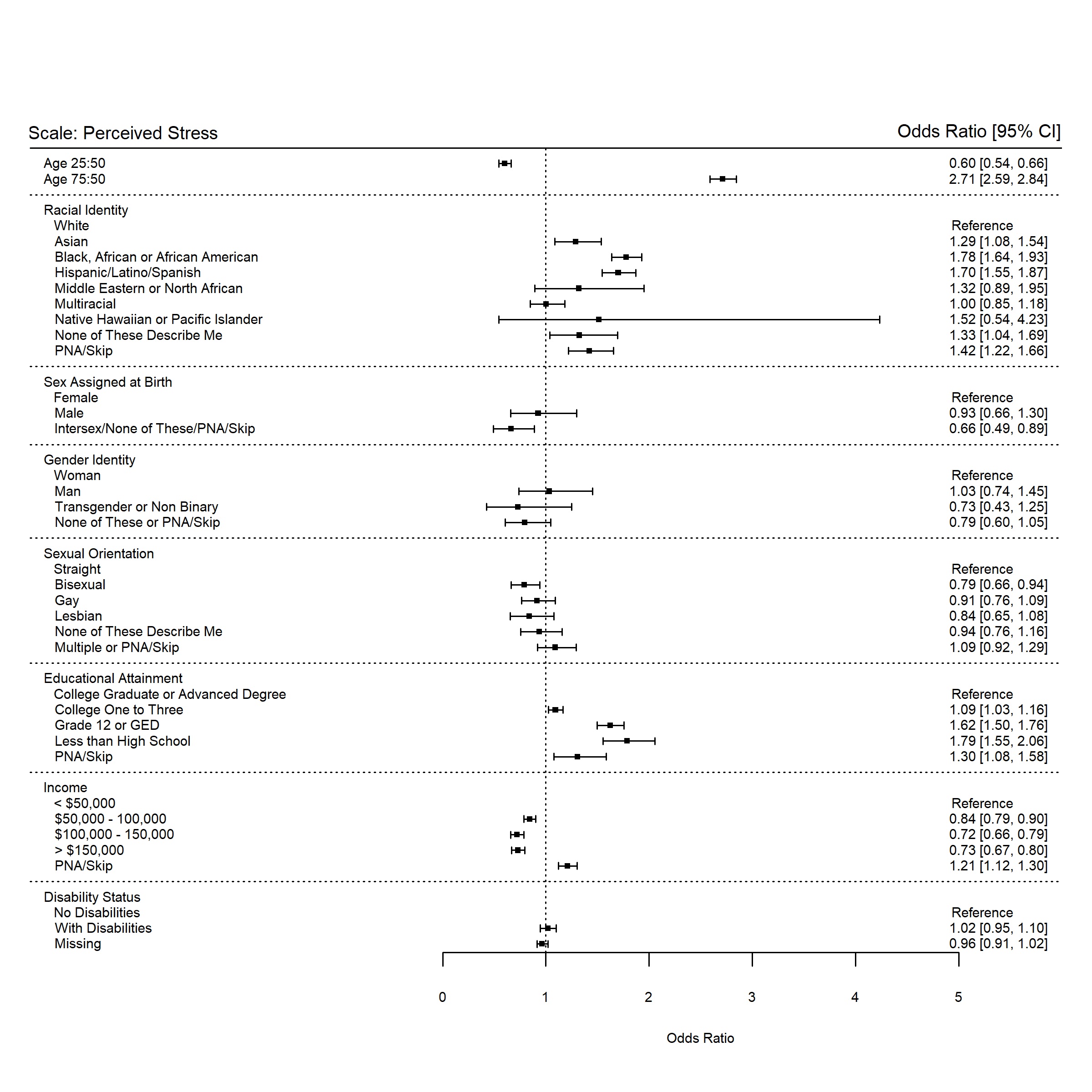

Notes: White; Black: Black, African or African-American; Hispanic: Hispanic/Latino/Spanish;

MENA: Middle Eastern or North African; NHPI: Native Hawaiian or Pacific Islander; PNA: Prefer not to answer

Sex at Birth: Sex Assigned at Birth

**Figure A8: Odds of item non-response or incalculable score for the Total Social Support Scale.**

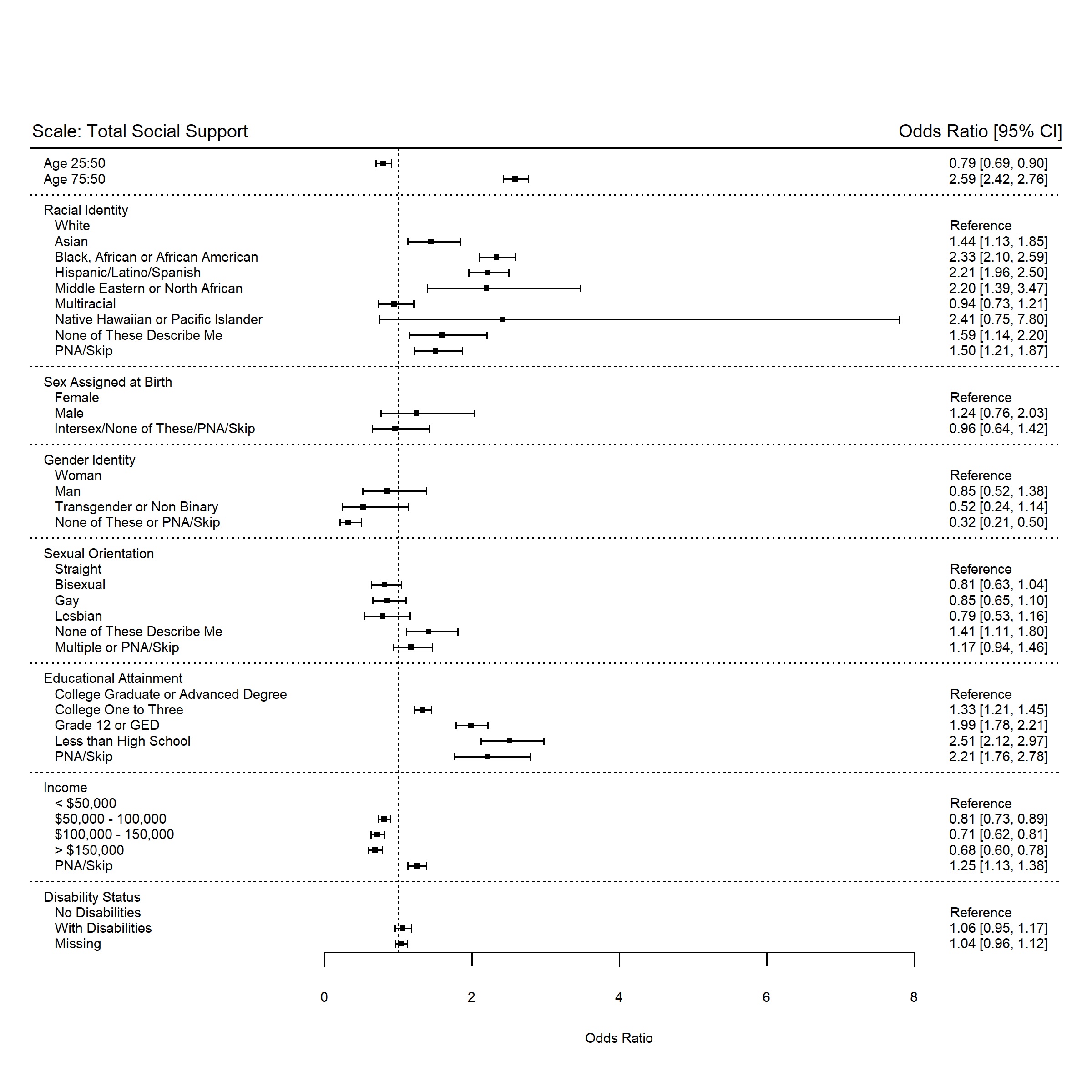

Notes: White; Black: Black, African or African-American; Hispanic: Hispanic/Latino/Spanish;

MENA: Middle Eastern or North African; NHPI: Native Hawaiian or Pacific Islander; PNA: Prefer not to answer

Sex at Birth: Sex Assigned at Birth

**Figure A9: Odds of item non-response or incalculable score for the Instrumental Social Support Scale**

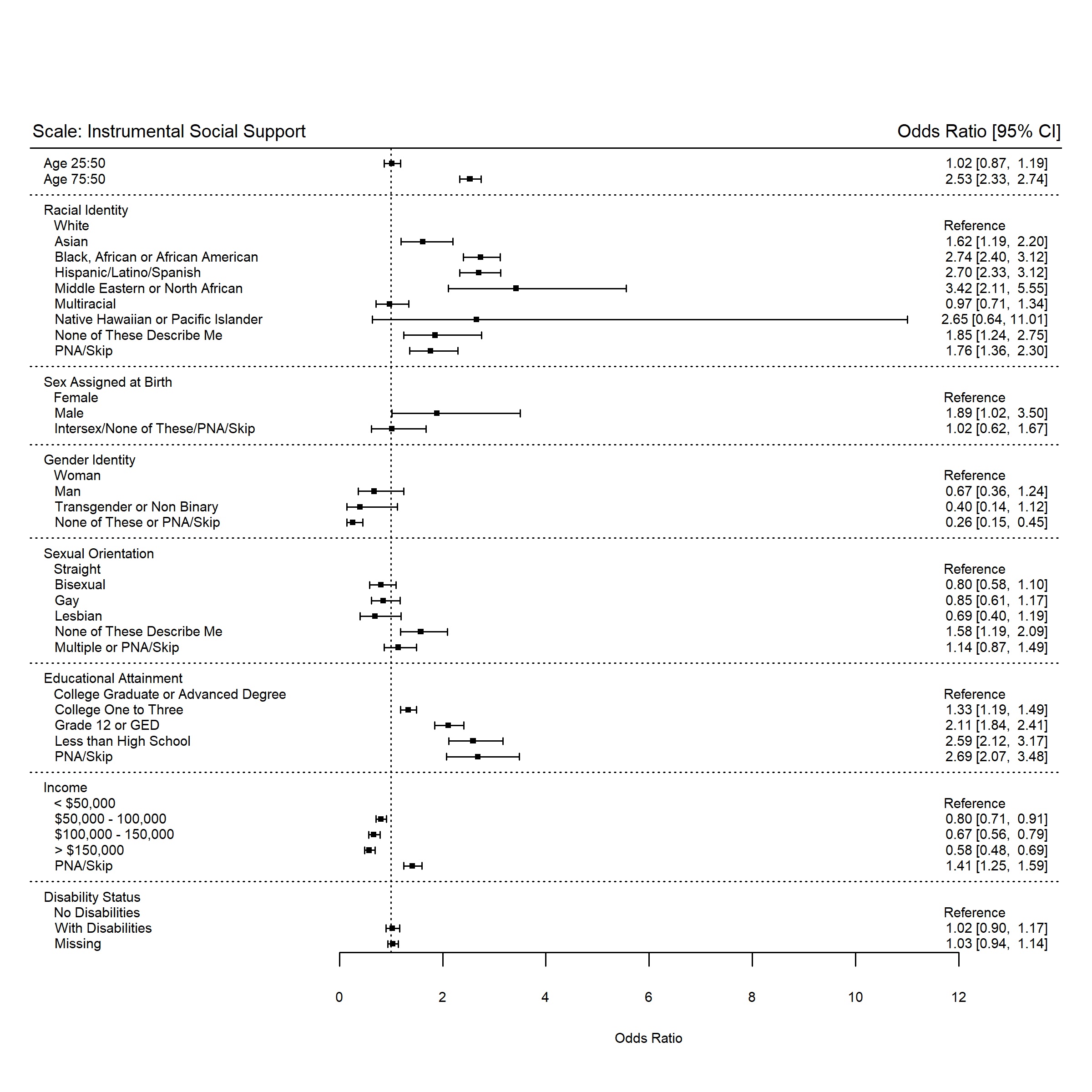
.

Notes: White; Black: Black, African or African-American; Hispanic: Hispanic/Latino/Spanish;

MENA: Middle Eastern or North African; NHPI: Native Hawaiian or Pacific Islander; PNA: Prefer not to answer

Sex at Birth: Sex Assigned at Birth

**Figure A10: Odds of item non-response or incalculable score for the Emotional Social Support Scale.**

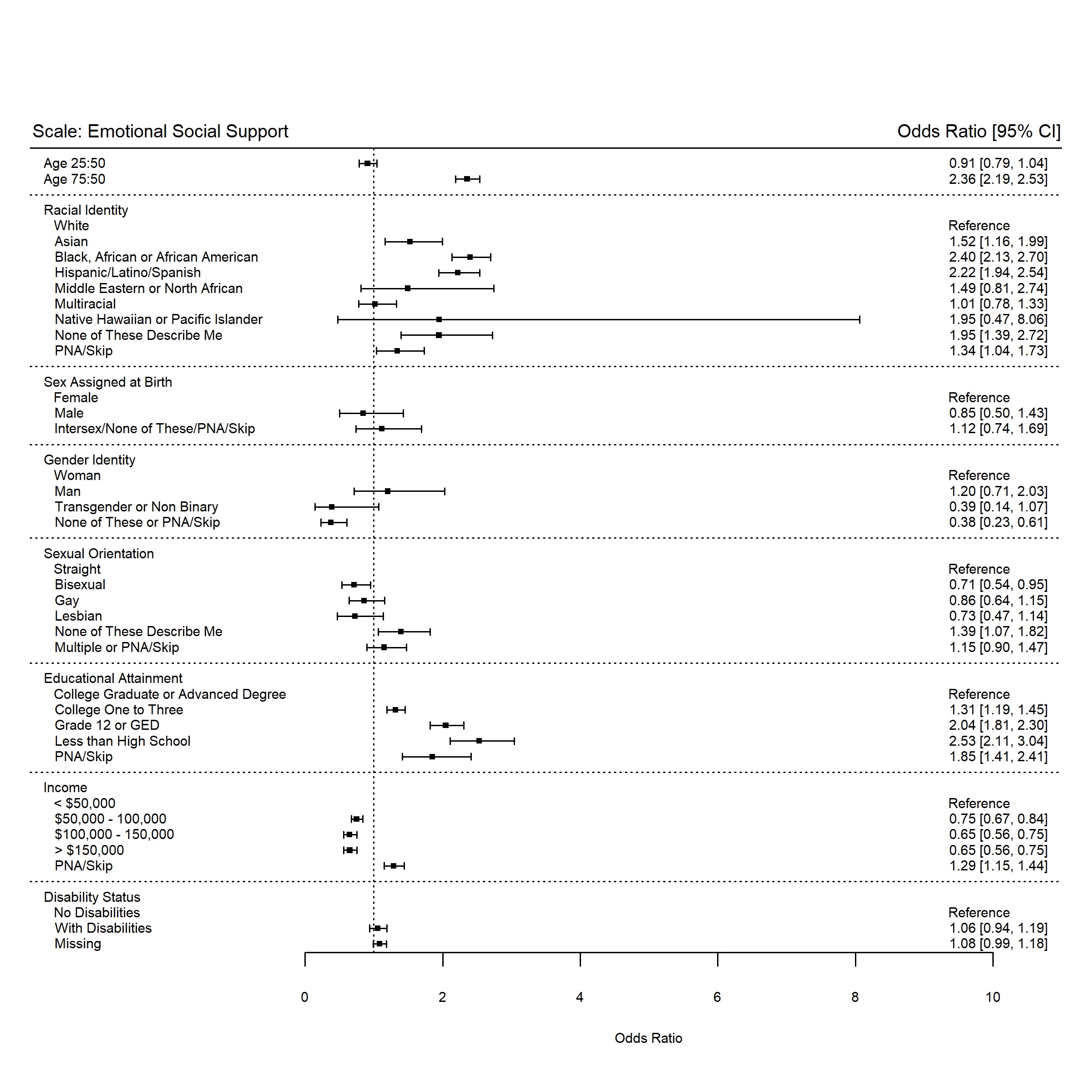

Notes: White; Black: Black, African or African-American; Hispanic: Hispanic/Latino/Spanish;

MENA: Middle Eastern or North African; NHPI: Native Hawaiian or Pacific Islander; PNA: Prefer not to answer

Sex at Birth: Sex Assigned at Birth

**Figure A11: Odds of item non-response or incalculable score for the Daily Spiritual Experiences Scale.**

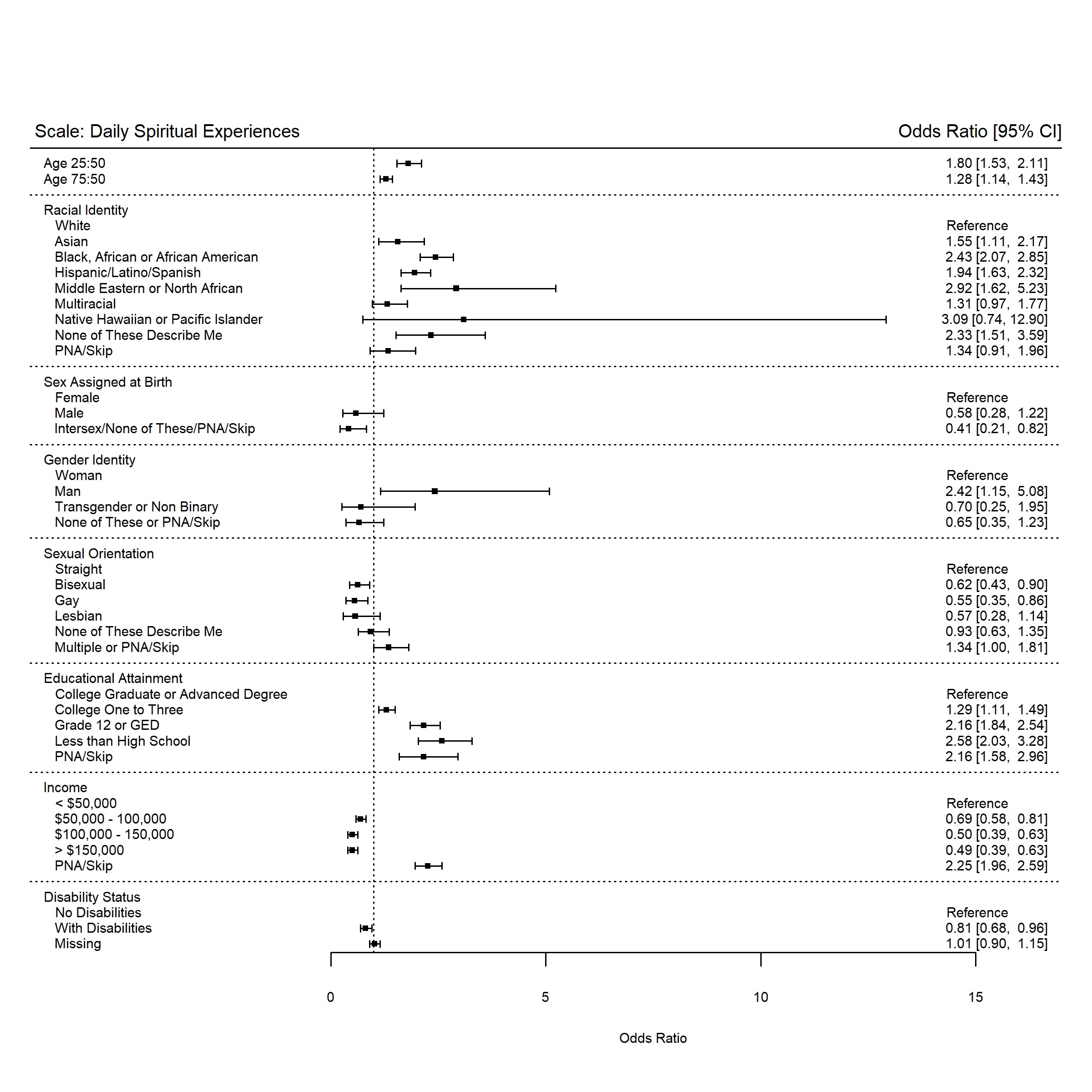

Notes: White; Black: Black, African or African-American; Hispanic: Hispanic/Latino/Spanish;

MENA: Middle Eastern or North African; NHPI: Native Hawaiian or Pacific Islander; PNA: Prefer not to answer

Sex at Birth: Sex Assigned at Birth

**Figure A12: Odds of item non-response or incalculable score for the Housing Instability Scale.**

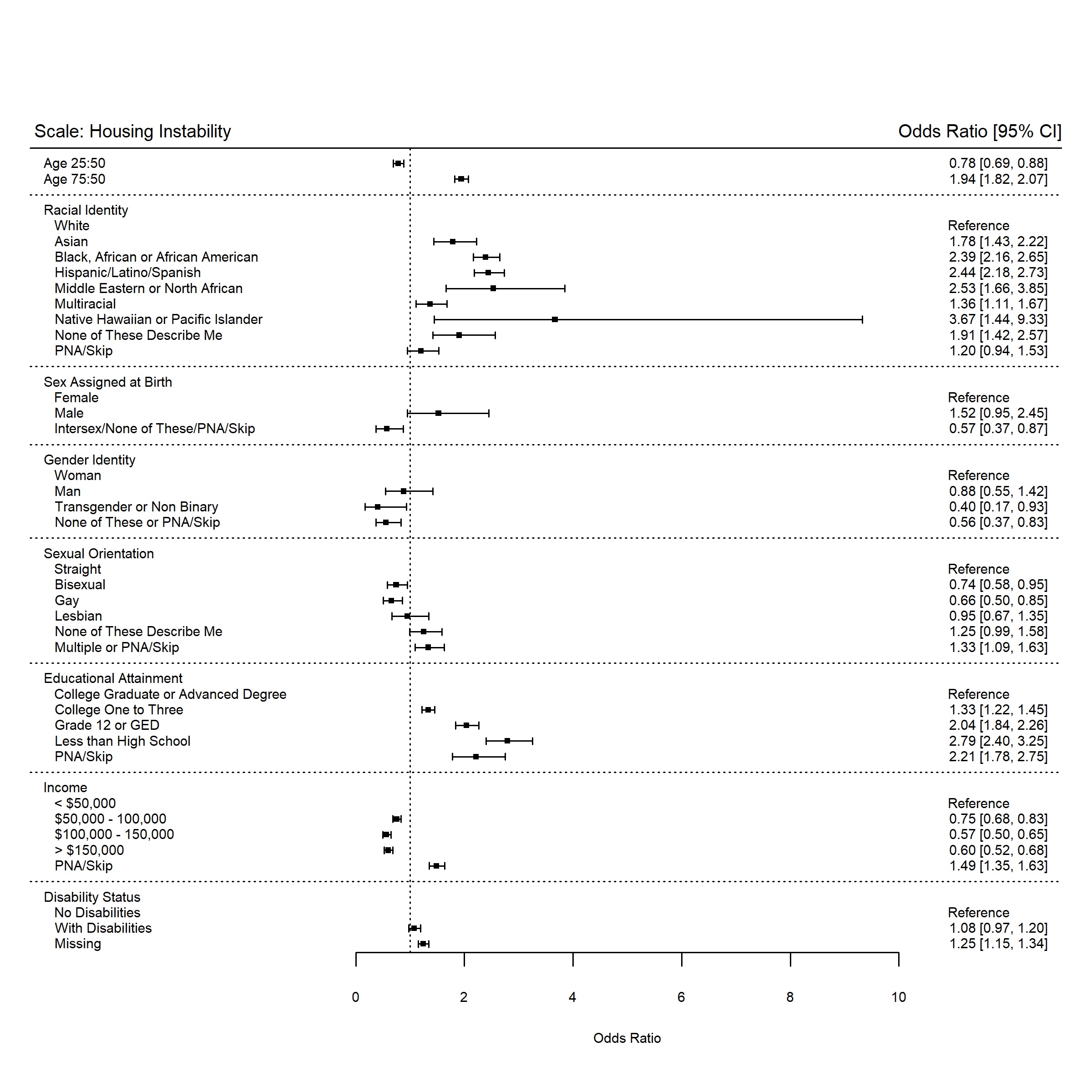

Notes: White; Black: Black, African or African-American; Hispanic: Hispanic/Latino/Spanish;

MENA: Middle Eastern or North African; NHPI: Native Hawaiian or Pacific Islander; PNA: Prefer not to answer

Sex at Birth: Sex Assigned at Birth

**Figure A13: Odds of item non-response or incalculable score for the Food Insecurity Scale.**

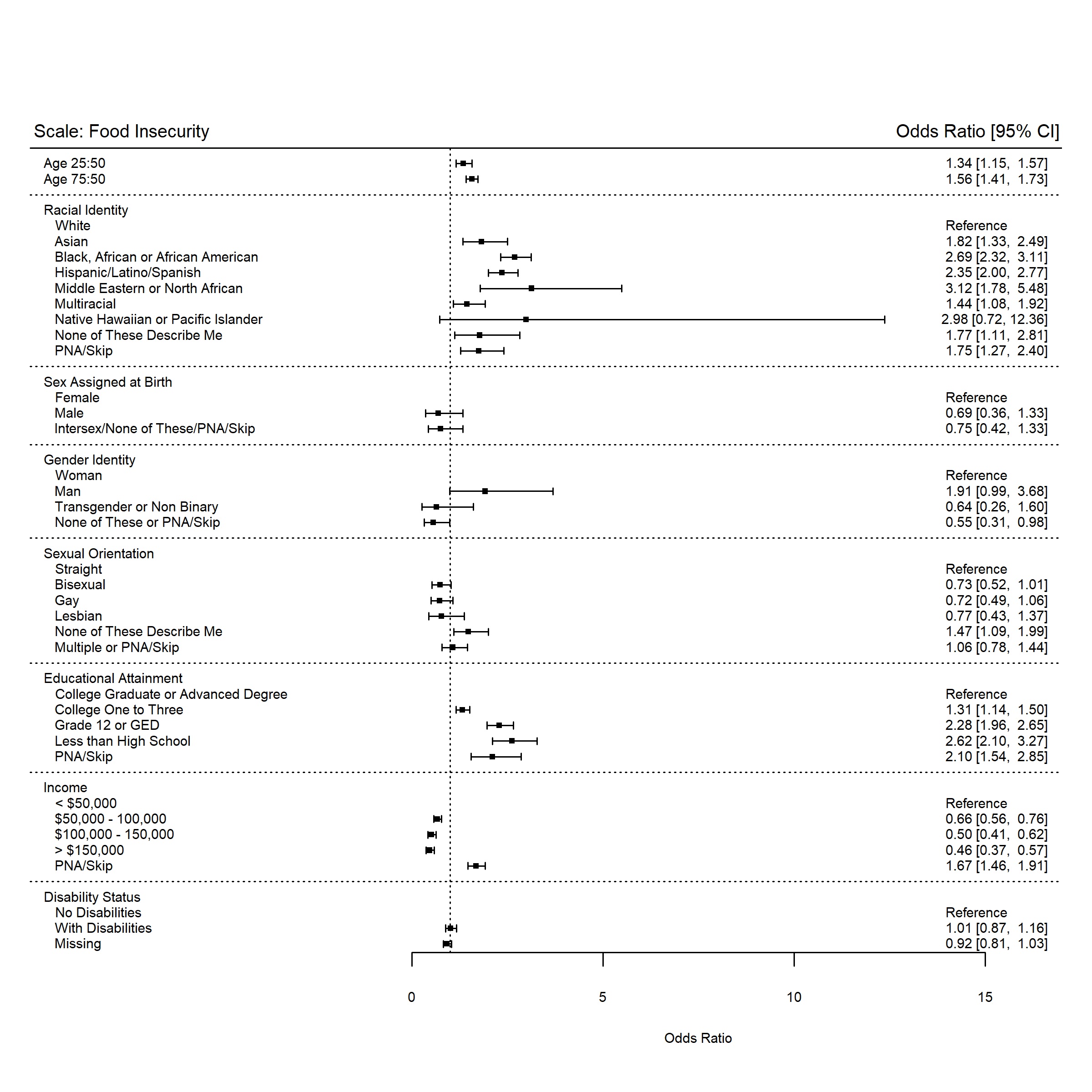

Notes: White; Black: Black, African or African-American; Hispanic: Hispanic/Latino/Spanish;

MENA: Middle Eastern or North African; NHPI: Native Hawaiian or Pacific Islander; PNA: Prefer not to answer

Sex at Birth: Sex Assigned at Birth

**Figure A14: Odds of item non-response or incalculable score for the PANES - Walking & Biking Scale.**

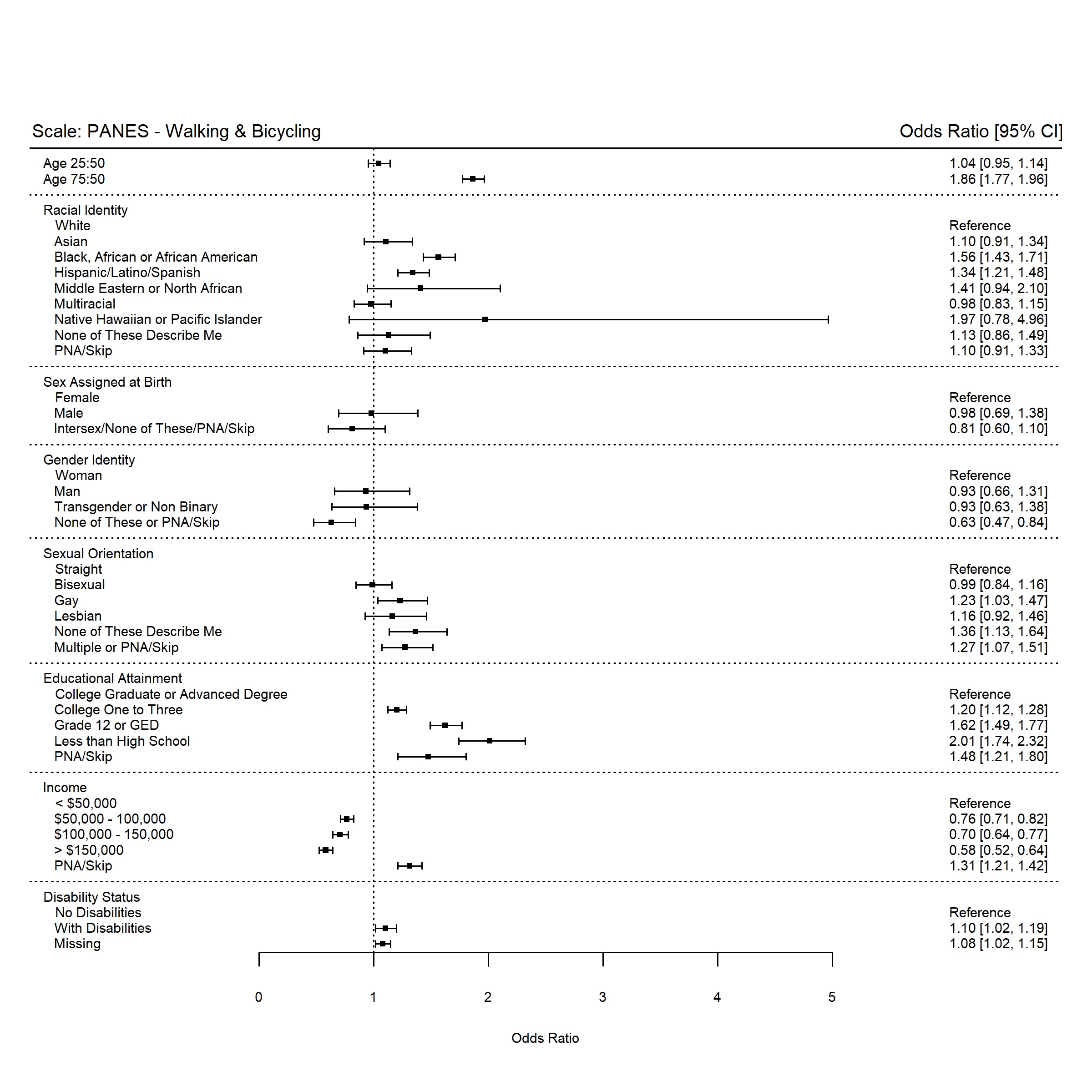

Notes: PANES: Physical Activity and Neighborhood Environment Scale; White; Black: Black, African or African-American; Hispanic: Hispanic/Latino/Spanish; MENA: Middle Eastern or North African; NHPI: Native Hawaiian or Pacific Islander; PNA: Prefer not to answer

Sex at Birth: Sex Assigned at Birth

**Figure A15: Odds of item non-response or incalculable score for the PANES - Crime & Safety Scale.**

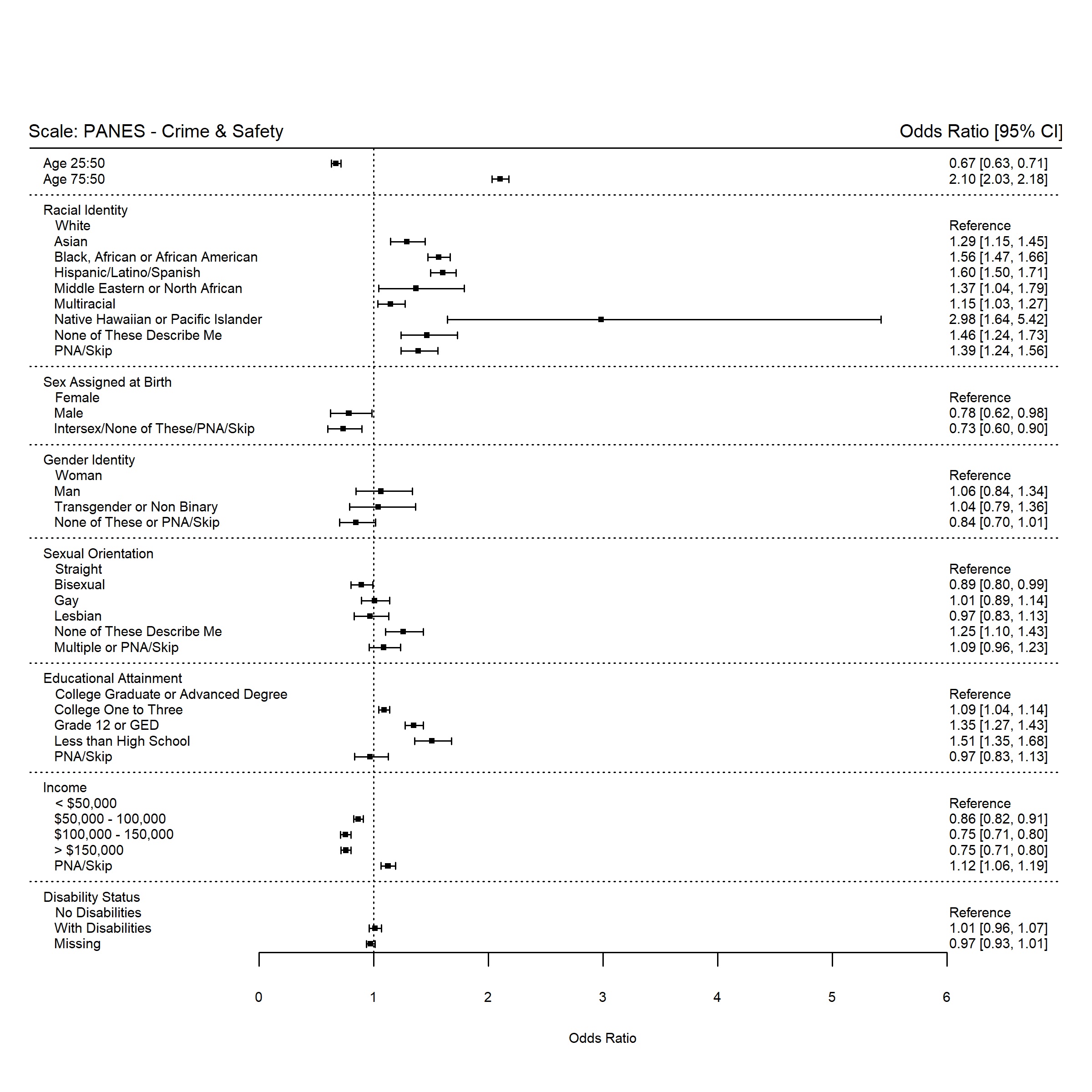

Notes: PANES: Physical Activity and Neighborhood Environment Scale; White; Black: Black, African or African-American; Hispanic: Hispanic/Latino/Spanish; MENA: Middle Eastern or North African; NHPI: Native Hawaiian or Pacific Islander; PNA: Prefer not to answer

Sex at Birth: Sex Assigned at Birth

**Figure A16: Odds of item non-response or incalculable score for the Housing Quality Scale.**

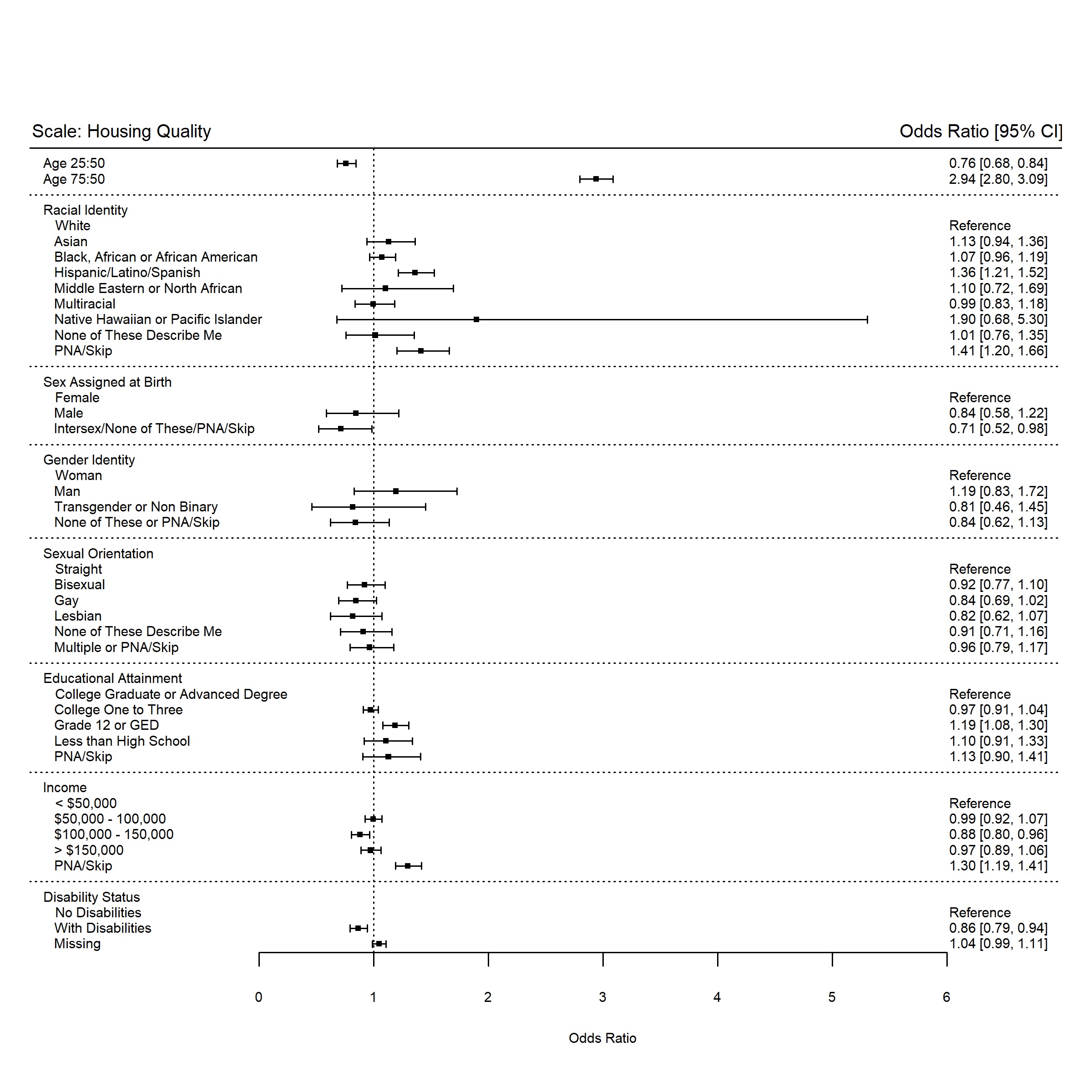

Notes: White; Black: Black, African or African-American; Hispanic: Hispanic/Latino/Spanish;

MENA: Middle Eastern or North African; NHPI: Native Hawaiian or Pacific Islander; PNA: Prefer not to answer;

Sex at Birth: Sex Assigned at Birth

**Figure A17: Odds of item non-response or incalculable score for the Religious Attendance Scale.**

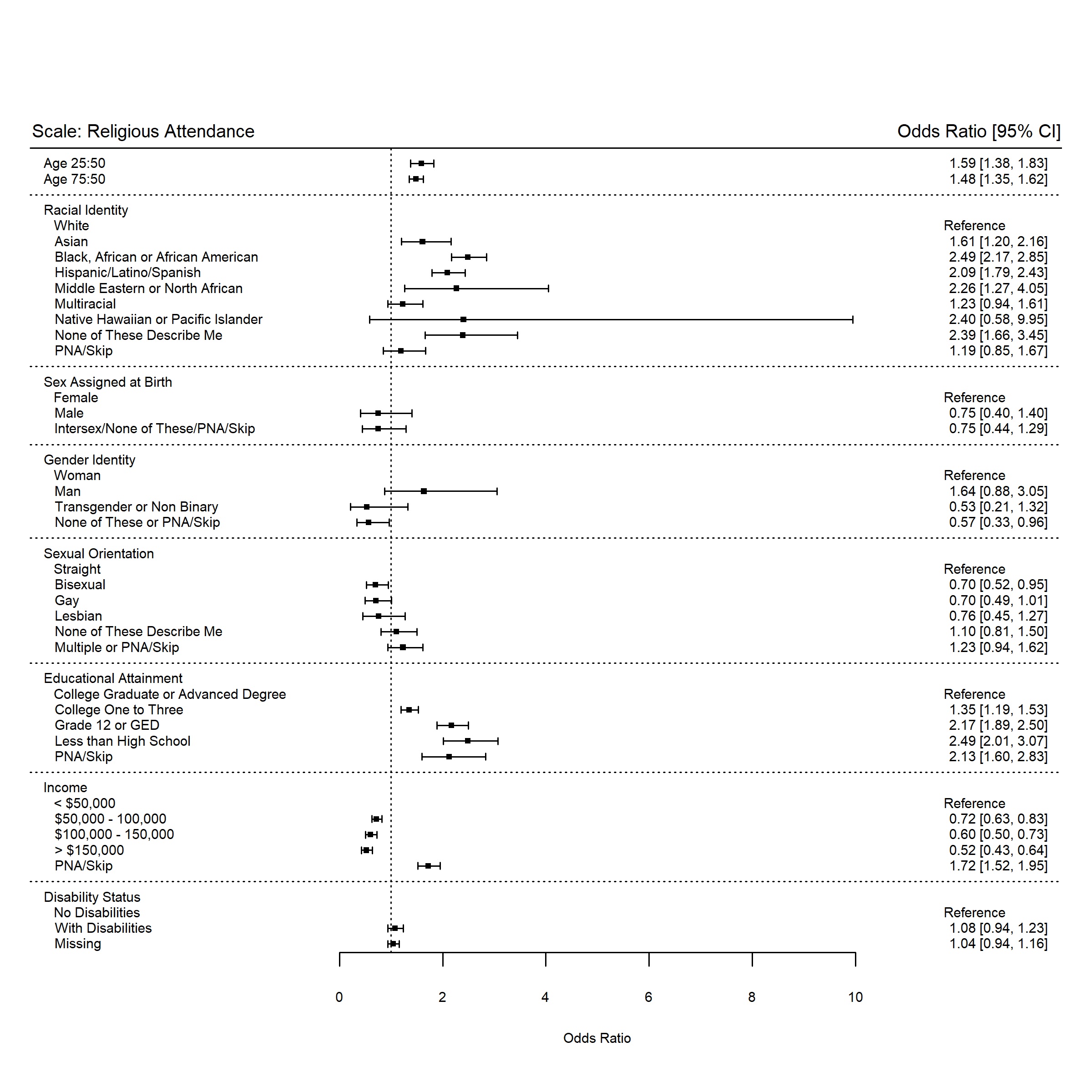

Notes: White; Black: Black, African or African-American; Hispanic: Hispanic/Latino/Spanish;

MENA: Middle Eastern or North African; NHPI: Native Hawaiian or Pacific Islander; PNA: Prefer not to answer

Sex at Birth: Sex Assigned at Birth
