## Appendix 6 for "Measuring Social Determinants of Health in the *All of Us* Research Program: Technical Document"

### Appendix 6: Acknowledgement List of Principal Investigators

**Past and Present *All of Us* Research Program Principal Investigators**

Brian Ahmedani^1^; Christine D Cole Johnson^1^; Habib Ahsan^2^; Hoda Anton-Culver^4^; Eric Topol^5^; Katie Baca-Motes^5^; Julia Moore-Vogel^5^; Praduman Jain^6^; Mark Begale^6^; Neeta Jain^6^; David Klein, MBA^6^; Scott Sutherland^6^; Bruce Korf^7^; Beth Lewis^7^; Ali G Gharavi^8^; George Hripcsak^8^; Eric Boerwinkle^9^; Scott Joseph Hebbring^10^; Elizabeth Burnside^11^; Dorothy Farrar-Edwards^11^; Amy Taylor^12^; Liliana Lombardi Desa^13^; Steve Thibodeau^14^; Mine Cicek^14^; Eric Schlueter^15^; Beverly Wilson Holmes^15^; Martha Daviglus^16^; Paul Harris^+17^; Consuelo Wilkins^17^; Dan Roden^17^; Kim Doheny^18^; Evan Eichler^19^; Gail Jarvik^19^; Gretchen Funk^20^; Anthony Philippakis^21^; Heidi Rehm^21^; Stacey Gabriel^21^; Richard Gibbs^22^; Edgar M Gil Rico^23^; David Glazer^24^; Jessica Burke^25^; Philip Greenland^26^; Elizabeth Shenkman^27^; William R Hogan^27^; Priscilla Igho-Pemu^28^; Elizabeth W Karlson^29^; Jordan Smoller^29^; Shawn N Murphy^29^; Cheryl R. Clark^51^; Margaret Elizabeth Ross^30^; Rainu Kaushal^30^; Eboni Winford^31^; Vik Kheterpal^32^; Francisco A Moreno^33^; Cheryl Thomas^34^; Mitchell Lunn^35^; Juno Obedin-Maliver^35^; Oscar Marroquin^36^; Shyam Visweswaran^36^; Steven Reis^36^; Patrick McGovern^37^; Gregory Talavera^38^; George T O'Connor^39^; Lucila Ohno-Machado^41^; Fornessa Randal^43^; Andreas A Theodorou^44^; Eric Reiman^44^; Mercedita Roxas-Murray^45^; Louisa Stark^46^; Ronnie Tepp^47^; Alicia Zhou^48^; Scott Topper^48^; Rhonda Trousdale^49^; Phil Tsao^50^; Scott T Weiss^51^; Jeffrey Whittle^53^; Stephan Zuchner^55^; Olveen Carrasquillo^55^; Megan Lewis^57^; Jen Uhrig^57^; May Okihiro^58^; Maria Argos^16^; Brisa Aschebook-Kilfoy^16^; Laura Bartlett^54^; Roberta Carlin^59^; Elizabeth Cohn^60^; Vivian Colon-Lopez^61^; Karl Cooper^59^; Linda Cottler^62^; Errol Crook^63^; Elizabeth Culler^64^; Charles Drum^59^; Milton Eder^62^; Mark Edmunds^52^; Rachel Everhart^65^; Adolph Falcon^23^; Becky Fein^66^; Zeno Frano^53^; Michael Garrett^67^; Sandra Halverson^68^; Eileen Handberg^27^; Joyce Ho^26^; Laura Horne^66^; Rosario Isasi^55^; Jessica Isom^69^; Jessica Jarmin^70^; Megan Jula^71^; Royan Kamyar^72^; Frida Kleiman^60^; Isaac Kohane^73^; Babbette Lamarca^67^; Brendan Lee^22^; Niall Lennon^21^; Dessie Levy^74^; Todd Mahr^75^; Emily Makahi^58^; Vivienne Marshall^76^;Elizabeth Mayer-Davis^77^; Jacob McCauley^55^; Jeffrey McKinney^78^; David McPherson^9^; Robert Meller^28^; Jose Melo^61^; David Ming-Hung Lin^79^; Michael Minor^74^; Evan Muse^5^; Kapil Parakh^80^; Cathryn Peltz-Rauchman^1^; Linda Rose Perez Laras^81^; Subhara Raveendran^82^; Gail Reilly^31^; Jody Reilly^83^; Nelida Rivera^81^; Laura Rosales^22^; Tracie Rosser^56^; Linda Salgin^38^; Sherilyn Sawyer^84^; William Simonson^85^; Amy Sitapati^41^; Cynthia So-Armah^69^; Gene Stegeman^86^; Christin Suver^87^; Michael Taitel^42^; Kyla Taylor^31^; Daniel Hernandez Tinoco^31^; Scott Topper^48^; Rhonda Trousdale^49^; Jason Vassy^84^; Jamie Walz^78^; Preston Watkins^88^; Blaker Wilkerson^89^; Katrina Yamazaki^12^; Melissa Basford^17^; Amrylis Silva Boschetti^41^; Suchitra Chandrasekaran^56^; Kim Enard^90^; Yuri Fresko^83^; Richard Grucza^90^; Robert Kelley^56^; Kathleen Keogh^13^; Cora Elizabeth Lewis^7^; Christopher Lough^91^; Ted Malmstrom^90^; David Ming-Hung Lin^79^; Paul Nemeskal^69^; Matt Pagel^56^; Jeffrey Scherrer^90^; Sanjay Shukla^10^; Debra Smith^92^; Bryce Turner^93^; Miriam Vos^56^

**Note**

This is the list of individuals who were Principal Investigators or equivalent with the *All of Us* Research Program during the period that this paper was in development, March 1, 2022 - June 1, 2023.

**Legend**

+ Principal Investigator/Lead Author for the *All of Us* Research Program protocol

**Affiliations**

1. Henry Ford Health System

2. University of Chicago Medical Center

3. Jackson-Hinds Comprehensive Health Center

4. University of California, Irvine

5. Scripps Research Translational Institute

6. Vibrent Health

7. University of Alabama at Birmingham

8. Columbia University

9. University of Texas Health Science Center at Houston

10. Marshfield Clinic Research Institute

11. University of Wisconsin at Madison

12. Community Health Center, Inc.

13. Sun River Health

14. Mayo Clinic and Foundation, Rochester

15. Cooperative Health

16. University of Illinois at Chicago

17. Vanderbilt University Medical Center

18. Johns Hopkins University School of Medicine

19. University of Washington

20. FiftyForward

21. Broad Institute

22. Baylor University

23. National Alliance for Hispanic Health

24. Verily Life Sciences

25. MITRE Corporation

26. Northwestern University

27. University of Florida

28. Morehouse School of Medicine, Atlanta

29. Partners Health Care

30. Cornell University, Weill Medical College

31. Cherokee Health Systems

32. CareEvolution, Inc.

33. University of Arizona, Tucson

34. Delta Research and Educational Foundation

35. Stanford University

36. University of Pittsburgh

37. Wondros

38. San Ysidro Health Center

39. Boston Medical Center

40. VA *All of Us* Coordinating Center, Boston

41. University of California, San Diego

42. Walgreen Co.

43. Asian Health Coalition

44. Banner Health

45. Montage Marketing Group

46. University of Utah

47. HCM Strategists

48. Color Genomics, Inc.

49. NYC Health + Hospitals

50. VA AoU Coordinating Center - Palo Alto

51. Brigham and Women's Hospital

52. San Diego Blood Bank

53. Medical College of Wisconsin

54. National Library of Medicine (NLM)

55. University of Miami School of Medicine

56. Emory University

57. Research Triangle Institute

58. Waianae Coast CHC

59. American Association of Health and Disability

60. Hunter College

61. University of Puerto Rico Comprehensive Cancer Center

62. CTSA Community Engagement Programs

63. University of South Alabama

64. TPC: Blood Assurance

65. TPC: Denver Health

66. TPC: Active Minds

67. University of Mississippi Medical Center

68. TPC: DLH Corp

69. Mass General Hospital

70. Tactis

71. TPC: Mary’s Center

72. TPC: Owaves

73. Harvard Medical School

74. National Baptist Convention

75. Gundersen Health System

76. South Texas Blood and Tissue Center

77. University of North Carolina at Chapel Hill

78. Sensis

79. TPC: Bloodworks Northwest

80. TPC: Fitbit

81. COSSMA

82. Patients Like Me

83. Quest Diagnostics Incorporated

84. VA AoU Coordinating Center

85. Cascade Regional BLood Services

86. ExamOne

87. Sage Bionetworks

88. WebMD Health Corp

89. Blue Cross Blue Shield

90. Saint Louis University

91. TPC: LifeSouth

92. TPC: SunCoast Blood Center

93. University of Southern California
